# Nigerian language as a contextual moderator of trauma-related PTSD and CPTSD among Liberian and Sierra Leonean refugee children in Oru Refugee Camp: A social-ecological moderation model of refugee children’s trauma

**DOI:** 10.64898/2026.03.11.26348168

**Authors:** Dogbahgen Alphonso Yarseah, Olu Francis Ibimiluyi, Ololade Omolayo Ogunsanmi, Omowumi Omojola Awosusi, J. Mac-Nixon Flomo, Bello Folaranmi Fatai, Elijah Olawale Olaoye, Alade Folasade Adesola

**Affiliations:** Ekiti State University, Faculty of Education, Department of Guidance and Counseling; Ekiti State University, Faculty of Education, Department of Guidance and Counseling Ado Ekiti, Nigeria.; Department of Public Health Babcock University, Ogun State, Nigeria.; Federal University of Oye-Ekiti Ekiti State, Nigeria Email.; University of Liberia, Amos C. Sawyer College of Social Sciences and Humanity, Department of Sociology and Anthropology & Criminology; Cavendish University of Uganda, Department of International Relations; Ekiti State University, Faculty Arts, Department of French Ekiti State, Nigeria; Ekiti State University, Facuty of Education, Department of Guidance and Counseling Ado Ekiti

## Abstract

**Background:** The closure of the Oru refugee camp in 2012 by the Nigerian government, following the withdrawal of legal and humanitarian support by the United Nations High Commission for Refugees, exposed Liberian and Sierra Leonean stateless refugee children to multiple and chronic traumatic experiences, resulting in forced relocation to an uninhabitable host community. To date, no study has examined their mental health outcomes. This study investigates how different trauma types affect complex PTSD (CPTSD), PTSD, and functional impairment, and examines whether Nigerian and parental language proficiency moderate these associations within a social ecological framework.

**Methods:** A cross-sectional study was conducted with 320 stateless refugee children aged 6–17 years (137 males, 183 females) in Nigeria. Trauma exposure and PTSD symptoms were assessed using DSM-5–based measures (CATS; CPSS-SR-5), while ICD-11 PTSD and CPTSD diagnoses were derived using the ITQ-CA. Functional impairment and school-based support were measured with standardized instruments. Data were analyzed using SPSS v22 and SmartPLS for moderation analyses.

**Results:** Based on ICD-11 criteria, 50.0% of participants met PTSD and 24.1% met CPTSD criteria. DSM-5 analyses indicated that 31.3% met full PTSD criteria, with many exhibiting subthreshold symptoms. Witnessed and physical trauma were strongly associated with PTSD severity and functional impairment, whereas emotional and sexual trauma were associated with disturbances in self-organization (DSO), indicative of CPTSD. Teacher support was associated with lower DSO symptoms; however, this effect was moderated by Nigerian language proficiency (β = −0.230, 95% CI [−0.338, −0.121]), such that support was protective only for children with higher Nigerian language proficiency.

**Conclusions:** Trauma-related psychopathology among stateless refugee children is highly prevalent and shaped by ecological and institutional conditions. School-based teacher support can protect against DSO symptoms, but its effectiveness depends on children’s linguistic access. These findings highlight the need to integrate ICD-11 CPTSD and DSM-5 PTSD frameworks and underscore the importance of linguistically inclusive, trauma-informed educational and mental health interventions in humanitarian settings.

## Introduction

### Background of the study

Protracted refugee crises constitute a persistent humanitarian challenge across sub-Saharan Africa, with children bearing a disproportionate share of the psychological burden associated with long-term displacement (UNHCR, 2024). Unlike short-term humanitarian emergencies, protracted displacement exposes children to repeated and cumulative stressors during critical developmental periods, increasing vulnerability to trauma-related mental health difficulties. In such contexts, children may no longer face immediate threats to physical survival, yet their psychological, educational, and social needs often remain unmet for extended periods, creating conditions of chronic adversity rather than acute trauma.

Sustained exposure to violence, loss, and instability is central to the construct of complex trauma, defined as repeated or prolonged traumatic experiences occurring in contexts where individuals have limited capacity to escape or seek protection (Herman, 1992). For refugee children, such exposure may involve witnessing violence, experiencing family separation, living in unsafe environments, and enduring prolonged uncertainty regarding legal status and future prospects. Trauma occurring under these conditions, particularly during childhood and adolescence, substantially elevates the risk of posttraumatic stress disorder (PTSD) and complex posttraumatic stress disorder (CPTSD) (WHO, 2018). However, despite the relevance of these frameworks, systematic examination of PTSD and CPTSD among stateless Liberian and Sierra Leonean refugee children remains limited.

In addition, intergenerational trauma research has largely focused on how psychological distress is transmitted from refugee parents to their children, often privileging adult legal status and resettlement outcomes. Such approaches, however, tend to overlook the everyday ecological environments in which refugee children experience, interpret, and manage trauma. For stateless refugee children, trauma is shaped not only by family histories but also by social, linguistic, and institutional conditions created by adult-centered humanitarian and policy decisions. These conditions may either intensify or buffer trauma-related PTSD and CPTSD outcomes. By situating children’s psychological outcomes within the social ecology of Oru Refugee Camp, the present study advances a contextually grounded social-ecological model of refugee children’s trauma that centers children’s lived realities in prolonged displacement.

The International Classification of Diseases (ICD-11) distinguishes PTSD and CPTSD as related but diagnostically distinct trauma-related disorders, a distinction particularly relevant for populations exposed to cumulative and prolonged adversity (Beckord et al., 2025). PTSD is characterized by re-experiencing, avoidance, and a persistent sense of threat, whereas CPTSD includes these core symptoms alongside broader disturbances in self-organization (DSO), encompassing affect dysregulation, negative self-concept, and interpersonal difficulties (Cloitre et al., 2013). Empirical evidence indicates that CPTSD is more strongly associated with chronic interpersonal trauma and is linked to more pervasive disruptions in emotional, social, and academic functioning than PTSD alone (Karatzias et al., 2017; Hyland et al., 2018).

Although growing empirical support for the ICD-11 PTSD–CPTSD distinction has emerged from studies conducted primarily in Europe and North America, research in West African refugee contexts remains sparse. This gap is particularly concerning given that the region has experienced sustained forced displacement over several decades. Available studies suggest that cumulative childhood trauma in refugee populations is associated with broader developmental and psychosocial difficulties, underscoring the importance of context-sensitive assessment frameworks that account for both symptom profiles and developmental processes (Hyland et al., 2018).

The distinction between PTSD and CPTSD is especially salient among Liberian and Sierra Leonean refugees residing in Nigeria, many of whom have experienced displacement extending over three decades. For children raised in these environments, trauma exposure has been compounded by prolonged uncertainty surrounding durable solutions, including repatriation, local integration, and third-country resettlement. Such uncertainty has been shown to undermine psychological stability, disrupt family systems, and intensify distress among refugee populations (Agblorti & Grant, 2019).

Despite the formal end of armed conflict in Sierra Leone (2002) and Liberia (2003), substantial numbers of refugees remained in Nigeria due to persistent political and economic instability in their countries of origin, limited repatriation support, unsuccessful resettlement applications, and failed local integration initiatives (Omata, 2012; Durodola, 2021). The closure of Oru Refugee Camp in 2012 further exacerbated these vulnerabilities. Following camp closure, formal refugee status was withdrawn, rendering many Liberian and Sierra Leonean families effectively stateless and without international protection. As a result, they were exposed to heightened discrimination, marginalization, and exclusion from host-community resources. The limited inclusion of displaced populations in host-country development planning is often linked to perceptions of refugees as economic liabilities or security threats, rather than as long-term residents with developmental needs (Durodola, 2018).

These structural conditions are consistent with processes of intergenerational trauma, whereby children are repeatedly exposed to emotional and physical violence, sexual victimization, and chronic insecurity within caregiving and community environments. Over time, refugee identity itself may become stigmatized, contributing to persistent stress exposure characterized by re-experiencing, emotional avoidance, and a heightened sense of threat. Such repeated violations of safety can have lasting consequences for emotional regulation, self-concept, and relational functioning.

The crisis confronting refugee children in Oru was further compounded by identity and belonging challenges. Many children had never visited their parents’ countries of origin, lacked proficiency in ancestral languages, and possessed limited cultural ties to Liberia or Sierra Leone.

At the same time, many adopted Nigerian languages through daily interaction, schooling, and social contact, yet linguistic adaptation did not necessarily translate into social acceptance within host communities. Developmental research indicates that sustained environmental instability and social exclusion can heighten children’s stress responsivity, weaken emotional regulation, and compromise long-term psychological well-being (Shonkoff, 2010; Shonkoff & Garner, 2012). Such conditions may elevate parental stress, which can undermine caregivers’ capacity to provide consistent emotional support, with downstream implications for children’s behavioral and socioemotional adjustment (Bronfenbrenner & Evans, 2000; Deater-Deckard et al., 2009; Evans & Kim, 2013)

Despite extensive research on trauma exposure and psychosocial support among refugee populations (Durodola, 2023 ;2021;Yarseah et al.,2025, Yarseah and Adegoroye, 2019), substantially less attention has been given to Liberian and Sierra Leonean Children areas of language as a contextual factor shaping trauma-related outcomes. Language functions not only as a medium of communication but also as a marker of social belonging, access to institutional resources, and the formation of supportive relationships with teachers and peers. In multilingual host settings such as Nigeria, proficiency in local languages may condition children’s ability to access social and educational support systems, potentially moderating the psychological impact of trauma exposure. However, empirical studies examining language use as a moderating factor in PTSD and CPTSD symptomatology among refugee children remain scarce.

Although research has examined intergenerational trauma and PTSD/CPTSD, it has largely focused on parents and symptoms, overlooking how children’s daily ecological environments shape the impact of trauma exposure. In protracted displacement settings like Oru Refugee Camp, social-ecological experiences such as physical, emotional, sexual, and witnessed trauma may interact with factors like linguistic integration, social support, and institutional interactions to influence PTSD and CPTSD outcomes. Yet these relationships remain underexplored. Addressing this gap is essential for developing child-centered, contextually informed interventions.

#### Statement of the Problem

Refugee children living in protracted displacement contexts are exposed to multiple, overlapping forms of trauma that extend beyond discrete war-related events. In addition to direct exposure to violence, many experience chronic stressors rooted in legal precarity, social exclusion, economic deprivation, disrupted education, and the withdrawal of institutional protection (Yılmaz et al., 2025; Marshall et al., 2016). These conditions create an ecological environment in which trauma exposure is cumulative, persistent, and developmentally embedded, placing refugee children at heightened risk for trauma-related psychopathology, including PTSD and CPTSD, as well as functional impairment (Bronfenbrenner, 1979; Betancourt et al., 2017).

While prior research has advanced understanding of intergenerational trauma and distinctions between PTSD and CPTSD, much of this work has remained parent-focused or symptom-centric, often overlooking the ecological environments in which refugee children live. Little is known about how children’s daily experiences—such as linguistic integration, interactions within schools and institutions, and access to parental and teacher support—shape the psychological impact of trauma. These gaps are particularly salient in protracted displacement settings like Oru Refugee Camp, where adult-centered policy and humanitarian decisions may restructure children’s ecological realities. Addressing this gap is critical for developing interventions and policies that are child-centered, contextually informed, and responsive to trauma-related outcomes.

Despite growing recognition of the ICD-11 distinction between PTSD and CPTSD (WHO, 2018), empirical research on these conditions among refugee children in West Africa remains limited. Most studies have focused on individual trauma exposure and PTSD symptom severity, often neglecting broader disturbances in self-organization (DSO)—including affect dysregulation, negative self-concept, and relational difficulties—that are core features of CPTSD (Cloitre et al., 2013; Brewin et al., 2017). This gap is particularly pronounced for stateless Liberian and Sierra Leonean children in Nigeria, many of whom have lived for decades under conditions of uncertainty and marginalization following camp closure and the withdrawal of international protection (Adepoju et al., 2010; UNHCR, 2022).

Refugee children living in protracted displacement contexts are exposed to multiple, overlapping forms of trauma that extend beyond discrete war-related events. In addition to direct exposure to violence, many experience chronic stressors related to legal precarity, social exclusion, economic deprivation, disrupted education, and the withdrawal of institutional protection (Yılmaz et al., 2025; Marshall et al., 2016). These conditions create an ecological environment in which trauma exposure is cumulative, persistent, and developmentally embedded, placing children at heightened risk for trauma-related psychopathology, including PTSD and CPTSD, as well as functional impairment (Bronfenbrenner, 1979; Betancourt et al., 2017).

While prior research has advanced understanding of intergenerational trauma and distinctions between PTSD and CPTSD, much of this work remains parent-focused or symptom-centric, often overlooking the ecological environments in which refugee children live. Little is known about how children’s daily experiences—such as linguistic integration, access to parental and teacher support, and interactions within schools and institutions—shape trauma-related outcomes. These gaps are particularly salient in settings like Oru Refugee Camp, where adult-centered policy and humanitarian decisions may restructure children’s ecological realities. Addressing these gaps is critical for developing interventions that are child-centered, contextually informed, and responsive to both PTSD and CPTSD outcomes.

Empirical research highlights the importance of family and teacher support in buffering the psychological impact of trauma (Betancourt et al., 2015; Alisic et al., 2012). Teacher support, however, is often limited by uncertainty and a lack of trauma-informed practices, even in high-exposure settings. Language proficiency is similarly under-theorized in ecological models of trauma, despite its central role in accessing education, psychosocial support, and social belonging. Evidence from other refugee populations indicates that greater host- and heritage-language integration is associated with reduced trauma-related symptoms, suggesting that linguistic context may function as a moderator of trauma outcomes (Montemitro et al., 2021; Kartal et al., 2019; Mavisakalyan et al., 2025).

Guided by the ICD-11 distinction between PTSD and CPTSD (WHO, 2018) and a social-ecological framework, the present study examines how social-ecological experiences—such as physical, emotional, sexual, and witnessed trauma—impact PTSD and CPTSD outcomes among stateless Liberian and Sierra Leonean refugee children. The study explicitly investigates parental and teacher social support as mediators of these relationships, while linguistic integration is examined as a moderator of the trauma–mental health pathways. By integrating trauma exposure, social support pathways, and language context, the study addresses critical gaps and advances a contextually grounded understanding of trauma-related mental health among displaced children in West Africa.

#### Objectives of the study

This study examines how trauma exposure—including witnessing, physical, emotional, and sexual trauma—affects PTSD symptoms, disturbances in self-organization (DSO), and functional impairment among Liberian and Sierra Leonean refugee children in Oru Refugee Camp. It investigates the mediating roles of parental and teacher support in these relationships and explores whether child Nigerian language use and parental language proficiency moderate the impact of trauma and social support on psychological outcomes. By addressing these objectives, the study aims to identify factors that exacerbate or buffer trauma-related symptoms, providing evidence to inform culturally and linguistically responsive interventions for refugee children.

#### Social ecological model of refugee children trauma

Figure 1 presents the Social-Ecological Model of Refugee Children’s Trauma, which provides a framework for understanding how multiple layers of influence shape trauma adjustment. At the structural and community level, factors such as colonial language marginalization, camp closure, statelessness, economic insecurity, and social exclusion create conditions of vulnerability. At the institutional and interpersonal level, teacher support, family support, and the language of schooling provide protective or risk pathways. At the individual level, trauma exposure interacts with child Nigerian language proficiency, which is hypothesized to moderate the impact of trauma on outcomes. Trauma-related outcomes include PTSD symptoms, disturbances in self-organization (DSO), and functional impairment. This model visually represents the study’s conceptual framework and guides the formulation of its objectives and hypotheses.

**Figure 1.**
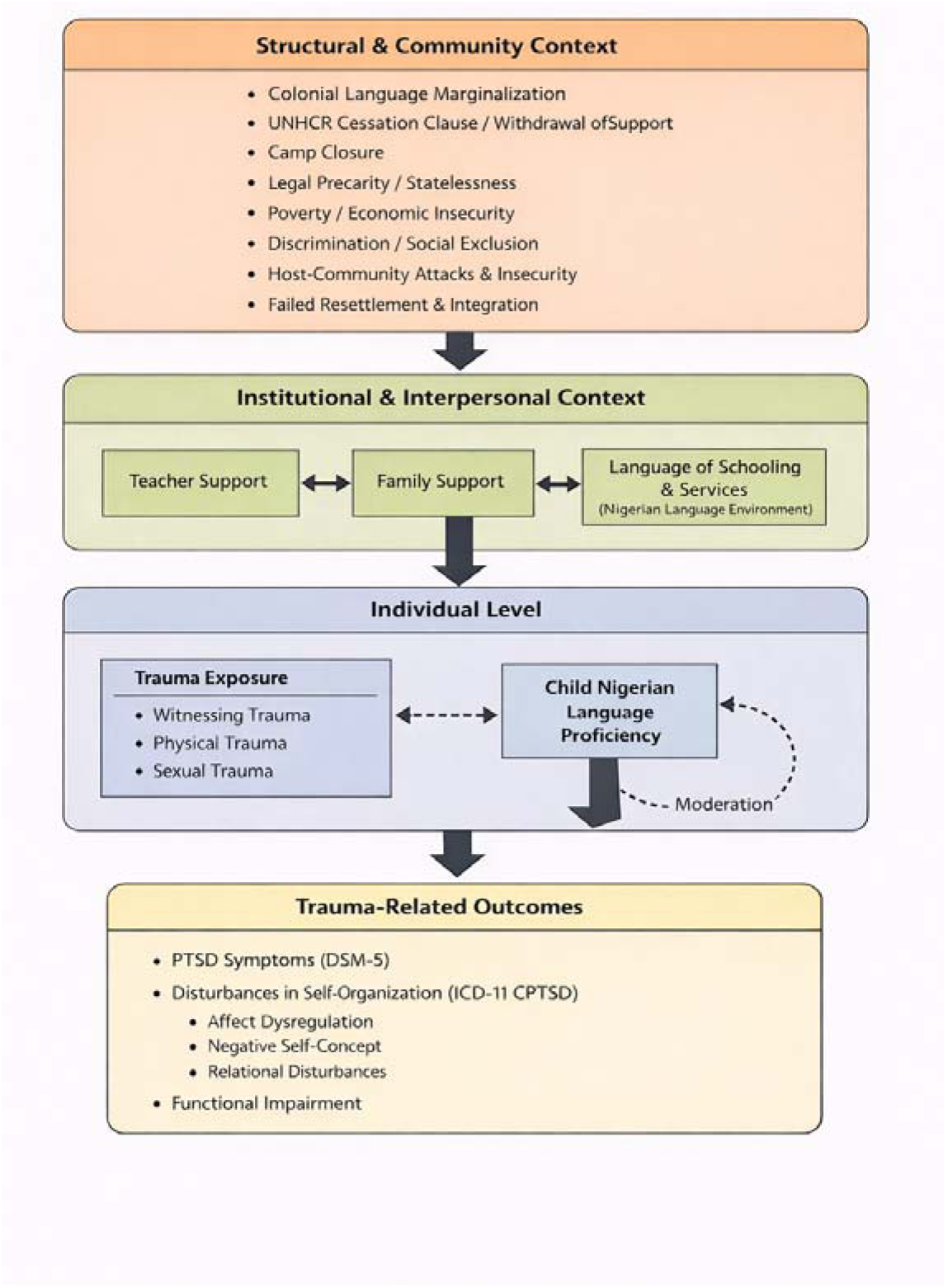
Social ecological framework of trauma-related psychopathology among stateless refugee children. Trauma exposure at the individual level interacts with gendered interpersonal experiences and institutional resources such as teacher support and language access. PTSD symptoms (DSM-5) are widespread and closely linked to physical and witnessed trauma, whereas CPTSD symptoms (ICD-11 disturbances in self-organization) emerge more strongly in the context of emotional and sexual trauma. Language proficiency moderates the protective effect of teacher support on disturbances in self-organization, illustrating how contextual resources shape trauma outcomes.

Although the present framework is informed by ecological thinking, it is not intended as a restatement of Bronfenbrenner’s ecological systems theory. Rather, it adapts a social ecological approach to the specific structural, institutional, and individual conditions shaping trauma exposure and posttraumatic outcomes among stateless refugee children. Unlike general developmental models, this framework explicitly incorporates ICD-11 distinctions between PTSD and complex PTSD, prolonged humanitarian exclusion, language marginalization, and post-camp displacement, thereby offering a contextually grounded model of trauma production rather than a general theory of child development.

The Social Ecological Framework is conceptualized in this study as a model for understanding refugee children’s trauma as the cumulative and interactive effects of multiple, nested environmental systems that shape psychological development over time. Rather than viewing trauma as the result of discrete or isolated events, this framework conceptualizes trauma as an ecological and developmental process emerging from sustained exposure to structural, environmental, relational, and individual-level adversities within the refugee camp context.

Refugee children’s psychological experiences often extend far beyond singular traumatic incidents and are deeply embedded within their social, political, and institutional environments. As Soguk (1999) argues, the refugee condition is characterized not only by the loss of home, nation, and citizenship, but also by the erosion of agency, voice, and social recognition what he describes as the absence of a “proper voice” and a “proper face.” For children growing up in stateless refugee camps, such losses are developmentally consequential, shaping early understandings of safety, belonging, identity, and the social world.

In examining trauma among refugee children in the fragmented environment of the abandoned Oru refugee camp, this study adopts a social ecological perspective to capture trauma exposure during a critical developmental period in which many children were born and raised. This period coincided with the withdrawal of UNHCR services, the closure of the camp by the Nigerian government, and forced relocation to degraded and unsanitary living conditions. Trauma is therefore conceptualized not as the outcome of discrete events, but as arising from the cumulative interaction of structural (policy neglect and service withdrawal), environmental (physical insecurity and deprivation), interpersonal (family disruption and sexual abuse), and intrapersonal (emotional and psychological distress) conditions within the camp.

The framework further proposes that the withdrawal of social services affects not only parents but also children’s early developmental environments, contributing to intergenerational patterns of trauma. Within this ecological context, children are exposed to multiple and overlapping adversities that jointly shape psychological functioning and long-term developmental outcomes.

Intrapersonal trauma refers to the internal psychological effects of traumatic exposure, including disruptions in emotional regulation, self-perception, and meaning-making systems (Chang et al., 2021). Among children, such trauma commonly manifests as heightened fear and anxiety, persistent emotional distress, and somatic complaints (Fazel & Stein, 2002). Children raised in chronically unstable and institutionally abandoned refugee camp environments may develop profound doubts about psychological safety, resulting in fragmented assumptions about themselves and the social world. Empirical studies further demonstrate that traumatic experiences in early childhood including abuse, neglect, and exposure to violence are associated with delays in physical, emotional, and social development (Jaycox et al., 2012), with accumulating evidence indicating adverse consequences for language acquisition and development (Hamidli, 2023), including reduced verbal competence, delayed expressive language, and lower speech intelligibility (Horsman, 2000).

In contrast, interpersonal trauma encompasses experiences of intentional harm, including physical, emotional, and sexual violence, as well as witnessing violence, which violate relational safety and disrupt psychological and social functioning. Evidence from a prospective birth cohort study indicates that exposure to interpersonal trauma—particularly during the first two years of life—is associated with significantly poorer cognitive functioning across early and middle childhood, even after controlling for socioeconomic and biological factors (Enlow et al., 2012).

Research indicates that during conflict and displacement, children frequently experience fears related to separation, abandonment, physical danger, and death (Moses et al., 2003). Such experiences undermine basic assumptions about safety and protection, heightening perceptions of vulnerability and disrupting coherence in children’s understanding of the world (Falsetti et al., 2003). From an intrapersonal perspective, these early traumatic exposures alter core belief systems and meaning-making processes, fostering negative worldviews characterized by low self-worth and perceptions of the world as unsafe or malevolent (Alsancak-Akbulut & Barişkin, 2020). These disruptions in self-concept and worldview represent key pathways linking early trauma exposure to later psychological outcomes, particularly disturbances in self-organization (DSO).

Within the social ecological framework of trauma, teachers occupy a critical role at the institutional level, directly influencing refugee children’s emotional and relational functioning. By providing co-regulation, predictability, and emotional validation, teachers can buffer the effects of intrapersonal trauma and support the development of adaptive coping and relational skills. However, working with trauma-exposed children can also place teachers at risk of secondary traumatic stress (STS) and compassion fatigue (CF), particularly in under-resourced educational settings (Hupe & Stevenson, 2019; Essary et al., 2020). Such strain may reduce their capacity to provide consistent emotional support, inadvertently limiting their protective influence on children’s intrapersonal functioning. These dynamics highlight that institutional supports are not only crucial for children’s resilience but also for maintaining teachers’ wellbeing, underscoring the importance of trauma-informed training, supervision, and adequate resources in refugee education contexts.

Tamm, & Tulviste, (2022) included children aged seven years and above, based on developmental evidence that by middle childhood core value structures are sufficiently established. Applying Schwartz’s theory of basic human values, which distinguishes ten universal motivational values (e.g., self-direction, benevolence, security), they identified a consistent value structure in children from age seven across diverse cultural contexts, suggesting the presence of sufficiently stable intrapersonal meaning systems (Tamm, & Tulviste, 2022; Döring et al., 2015; Cieciuch et al. 2016). Because intrapersonal trauma involves disruptions to internal beliefs, values, and meaning-making processes, the presence of these organized value structures constitutes a necessary precondition for examining trauma pathways and the buffering role of institutional supports such as teachers. By focusing on this developmental stage, the study ensures that both intrapersonal vulnerabilities and social ecological interventions can be meaningfully assessed.

#### Risk and Resilience Perspective of Stateless Refugee Children

Research on risk and resilience has evolved from early deficit-oriented models focused on pathology toward strengths-based perspectives that emphasize positive adaptation in the context of adversity (VicHealth; 2015). Risk refers to exposure to adverse life conditions—such as poverty, family discord, parental illness, violence, or trauma—that increase vulnerability to maladaptive developmental outcomes (Werner;1982). Importantly, risks rarely occur in isolation; rather, they tend to accumulate and cluster, exerting compounded effects on psychological functioning in a dose–response manner (Garmezy; 1991; Masten; 2001;2011; 2014). For Liberian and Sierra Leonean refugee children residing in Nigeria, displacement represents not a single stressor but a convergence of multiple, intersecting risks, including statelessness, chronic poverty, trauma exposure, disrupted education, and language marginalization, all of which jointly heighten vulnerability to adverse mental health outcomes.

Resilience, in contrast, is not a fixed trait but a dynamic and developmental process shaped by the interaction between risk exposure and protective resources. Rutter(2006;2013) conceptualized resilience as positive adaptation despite significant adversity, emphasizing that competence can only be meaningfully interpreted in the presence of risk. Similarly, Garmezy (1987; 1991) described resilience as the balance between cumulative risks and available protective factors, whereby favorable outcomes become more likely when protective resources outweigh exposure to adversity. Luthar (2000) further reinforced this perspective, defining resilience as a process of adaptation that requires both demonstrable risk exposure and evidence of adaptive functioning.

A consistent theme across resilience theorists is that positive adaptation typically emerges through ordinary, everyday systems of support rather than extraordinary individual traits. Masten (2011;1991) described this phenomenon as “ordinary magic,” highlighting the protective role of basic adaptive systems such as supportive caregiving relationships, responsive schools, and stable social environments. Werner (1982) extended this perspective ecologically, demonstrating that protective processes often operate indirectly across multiple levels—for example, when external support to caregivers enhances their capacity to provide emotional security to children. Ungar (2005;2013) further emphasized that resilience is culturally and contextually embedded, depending not only on individual coping strategies but also on access to institutional, relational, and material resources.

Importantly, these risk and resilience perspectives do not constitute the primary theoretical framework of the present study, but instead function as an explanatory lens within the broader Social Ecological Framework. Within this ecological context, family and teacher support are conceptualized as key protective processes that may mediate or buffer the effects of cumulative trauma exposure on psychological outcomes, including posttraumatic stress disorder (PTSD), complex PTSD (CPTSD), disturbances in self-organization (DSO), and trauma-related functional impairment. For stateless refugee children in Nigeria—whose protective environments have been significantly eroded by prolonged camp closure, humanitarian withdrawal, and social exclusion—examining how family and teacher support operate as resilience-building processes provides critical insight into mechanisms of adaptation and informs the development of contextually grounded interventions.

### Trauma Exposure and Mental Health

The closure of the refugee camp, displacement into unstructured urban environments, denial of resettlement, and statelessness without reliable governmental or international support constitute profound risk factors for refugee children. In Masten’s framework, such conditions represent high adversity coupled with scarce protective resources, a dynamic she identifies as central to vulnerability(Masten;2014; 1999). The United Nations Refugee Agency does not list Liberia and Sierra Leone in its classification of refugees, and they are rarely mentioned in UNHCR reports, as their category is often treated as a “finished programme”(Omata;2014). The withdrawal of humanitarian support, however, has been described as equivalent to the termination of life, as it exposes refugees to extreme vulnerability(Omata;2014). The 2012 cessation clause therefore served as a major trigger of trauma exposure among refugee children, which has long been associated with adverse mental health outcomes.

Childhood trauma—including witnessing violence, or experiencing physical, emotional, or sexual abuse—has been strongly linked to later mental health challenges. According to the World Health Organization, mental health is a state of wellbeing that enables individuals to cope with the stresses of life, realize their abilities, learn, work productively, and contribute to their communities (WHO; 2022). Many refugee children in Oru were born during a period of socioeconomic withdrawal by the international community. Those children who demonstrated resilience under such conditions could be said to have relatively good mental health. As Werner (2005)] noted, “not all children succumb to adverse life events.”

Nevertheless, risk factors such as poverty, growing up in disadvantaged communities, prolonged family disruptions, homelessness, parental discord, or exposure to parental alcoholism and mental illness (Werner;1989), may heighten the likelihood of developing mental health disorders. These disorders have been defined as clinically significant disturbances in cognition, emotional regulation, or behavior, reflecting dysfunction in underlying psychological, biological, or developmental processes ((International Advisory Group for the Revision of ICD-10 Mental and Behavioural Disorders [IAG], 2011). People with severe mental disorders are at increased risk of premature death due to suicide and comorbid conditions such as cardiovascular disease, diabetes, infectious diseases, and undernutrition. Having a severe mental disorder is associated with a 60% higher likelihood of dying prematurely from a noncommunicable disease, and up to 8% of global years of life lost are attributed to excess deaths linked to mental health conditions(Vigo, Thornicroft &, Atun;2016).

Untreated mental health conditions in childhood and adolescence can also result in educational difficulties, poor physical health, and social exclusion, ultimately leading to worse health outcomes and lower economic productivity in adulthood. Studies further suggest that the prevalence of mental health conditions among displaced and refugee children resettled in high-income countries is higher than in their non-refugee peers (Fazel et, al; 2012).

Research has consistently revealed a direct relationship between trauma exposure and PTSD and language development among children [Dvir et, al; 2014; Dar, Iqbal &, Mushtaq; 2015; Li, Seng;2018; Shenk, Putnam &, Noll;2014). Notably, however, there is a paucity of research on the mental health challenges of refugee children in the Oru camp following the 2012 cessation clause—representing a critical gap this study seeks to address.

#### DSM-5 Post-Traumatic Stress Disorder among Refugee Children

Studies have shown that children living in refugee camps often experience significant mental health challenges, yet many of these studies have not specifically applied the DSM-5 criteria for PTSD. Investigating PTSD through the DSM-5 framework in the context of long-term displacement is crucial, as it ensures consistency in diagnosing trauma-related conditions and enhances comparability across research and clinical settings. While previous scholars have documented the psychological impact of prolonged stress on displaced children—highlighting increased risks of depression, anxiety, PTSD, and maladaptive coping strategies (Blackmore et al., 2020; European Society for Traumatic Stress Studies[ESSTSS];2023) the DSM-5 provides a refined classification system that distinguishes between PTSD and Complex PTSD (CPTSD).

According to the DSM-5, PTSD develops following exposure to actual or threatened death, serious injury, or violence. This exposure may occur through direct experience, witnessing traumatic events, learning that a traumatic event happened to a close friend or family member, or repeated exposure to distressing details of trauma (children encountering graphics information of their parents execution by armed men)(American Psychiatric Publishing; 2013[APA];2013). The DSM-5 defines PTSD across four symptom clusters—re-experiencing, avoidance, negative alterations in cognition and mood, and hyperarousal/reactivity—and introduces new symptoms such as persistent negative beliefs and reckless behavior (APA, 2023).

While research on the DSM-5 PTSD framework has been conducted in low-income countries, studies in African contexts, particularly among refugee children, remain limited (Michalopoulos et al.; 2015). No known studies have specifically examined PTSD symptomatology using DSM-5 criteria among West African refugee children who have endured protracted displacement in the Oru refugee camp for over 18 years. Given the high prevalence of trauma exposure in this population such as witnessing trauma, emotional and sexual trauma, assessing PTSD symptoms through the DSM-5 framework is critical since we have no empirical literature that have identified PTSD symptomology among this population.

The DSM-5 also acknowledges developmental differences in trauma responses. Although Complex PTSD (CPTSD) and Developmental Trauma Disorder (DTD) were not included as formal diagnoses, the DSM-5 introduced a PTSD specifier for children under six to account for developmental variations (Resick et al., 2012; Friedman, 2013). For refugee children, prolonged trauma exposure may lead to emotional dysregulation, cognitive difficulties, interpersonal struggles, and identity disturbances—symptoms often associated with CPTSD rather than PTSD alone.

#### Complex PTSD in Children

Complex PTSD (CPTSD) was formally introduced in ICD-11 (World Health Organization, 2019) following earlier conceptualizations of disorders of extreme stress not otherwise specified (DESNOS) (van der Kolk et al.; 2005). Unlike PTSD, which is typically linked to single traumatic events such as accidents or combat, CPTSD arises from repeated, prolonged, and relational trauma(Brewin, 2020). It combines the core PTSD features of re-experiencing, avoidance, and sense of threat with disturbances in self-organization (DSO), including affect dysregulation, negative self-concept, and relational difficulties (Brewin, 2020). These DSO symptoms highlight the broader psychosocial impact of complex trauma beyond PTSD alone (Maercker et al.; 20222).

ICD-11 allows CPTSD diagnosis across the lifespan, including in children and adolescents, and emphasizes that children may be at particularly high risk due to developmental vulnerability (WHO, 2018; Cloitre et al., 2019) such as parental displacement, environmental stress, witnessing trauma, malnutrition, stunting, inadequate psychological stimulation, poverty disruption in emotional regulation, and linguistic development due to the cumulative effects of trauma, displacement, and deprivation. Evidence shows that CPTSD is more likely to result from childhood trauma than PTSD, with greater social dysfunction and cumulative adversity (Karatzias, et al., 2017). This makes CPTSD especially relevant for refugee children who have endured prolonged displacement, disrupted caregiving, and ongoing instability, as seen in Oru.

Compared with DSM-5, which requires a larger set of symptom clusters, ICD-11 simplifies PTSD to six symptoms in three clusters, while CPTSD adds DSO as a distinct dimension(Sarr et al., 2024). This framework has proven clinically useful in differentiating simple from complex trauma presentations in young populations.

Empirical studies outside Africa provide prevalence estimates of CPTSD among adolescents: 3.4% in Ireland vs. 1.5% PTSD(Redican et al., 2022), 4.1% in Italy vs. 9.3% PTSD (Kazlauskas eta al., 2022) 12.3% in Lithuania vs. 5.2% PTSD (Rossi et al., 2022), and 4.1% in Japan vs. 2.3% PTSD (Kazlauskas eta al., 2022). Risk factors for CPTSD include multiple trauma exposures (particularly sexual trauma), school and family difficulties, and female gender (Daniunaiteet al., 2021; Redican et al., 2022). Longitudinal research also shows varied trajectories, with some adolescents developing chronic CPTSD and others recovering over time.

Despite these findings, there are no empirical records of CPTSD among West African refugee children, particularly those growing up in disbanded camps like Oru without international support. Understanding CPTSD in this population is critical to tailoring interventions that address the unique developmental and psychosocial effects of prolonged trauma in stateless refugee children

#### Witnessing Trauma, and Its Impact on PTSD, CPTSD and Functional impairment

The effects of witnessing trauma on children’s mental health, particularly its association with PTSD and CPTSD, have been widely documented. Research indicates that children who witness traumatic events are at significant risk of developing PTSD symptoms, including re-experiencing, avoidance, and hyperarousal. For example, in a study of children aged 7–11 who witnessed the execution of a criminal near their school, 52% exhibited PTSD symptoms: 88 children experienced re-experiencing, 24 displayed avoidance, and 62 reported hyperarousal (Attari, Dashty & Mahmood 2006). A limitation of this study, however, was the use of the PTSD Civilian Checklist developed for adults rather than an age-appropriate tool for children.

Recognition of developmental differences has been central in subsequent diagnostic refinements. The DSM-IV emphasized that PTSD in children often manifests through play reenactment or frightening dreams rather than verbalized distress (Scheeringa et al, 1995). Later studies introduced developmentally sensitive criteria for preschool-aged children, incorporating symptoms such as aggression, new fears, and regression in previously acquired developmental skills, including language (Margolin, & Vicerman, 2010).

A more recent large-scale study in the Gaza Strip examined both personal trauma and witnessing trauma among children aged 7–17. Findings revealed that 88.4% experienced personal trauma, 83.7% witnessed trauma to others, and 88.3% observed property demolition. More than half (53.5%) met DSM-5 criteria for PTSD. Even after adjusting for demographic and socioeconomic variables, personal trauma, witnessing trauma, and property destruction all predicted PTSD, with personal trauma exerting the greatest effect, followed by witnessing trauma, and then property loss (El-Khodary, Samara & Askew 2020). These findings underscore that witnessing trauma is a consistent predictor of PTSD in children, although methodological considerations and developmental appropriateness remain critical.

Beyond clinical symptomatology, trauma-related disorders in children are strongly associated with impairments in daily functioning, academic achievement, and social relationships. Functional impairment is increasingly recognized as a critical outcome because it reflects the broader impact of trauma on children’s developmental trajectories and long-term well-being. Functional impairment is a critical but underexplored dimension of trauma-related psychopathology in children (Maercker et al., 2022). Population-based surveys and clinical studies consistently demonstrate that mental disorders are associated with significant impairments across domains of social, academic, and relational functioning(Cross et al., 2023). The diagnostic frameworks of both DSM-5 and ICD-11 underscore this point: PTSD and CPTSD diagnoses require that symptoms cause clinically significant distress or functional impairment, whether in school, peer relationships, or family life. Yet, impairment is not always present at diagnostic threshold, making it essential to examine the specific symptom clusters most predictive of functional difficulties.

In line with both ICD-11 and DSM-5 diagnostic frameworks, functional impairment is a central outcome of trauma-related disorders. The present study therefore employs three indicators of functional impairment: PTSD functional impairment (ICD-11), DSO functional impairment (ICD-11 CPTSD), and PTSD functional impairment (DSM-5). ICD-11 distinguishes between impairments linked to threat-based PTSD symptoms (intrusions, avoidance, current sense of threat) and those arising from disturbances in self-organization (DSO), such as affect dysregulation, negative self-concept, and interpersonal disturbances (Cloitre et al., 2018). This distinction is supported by evidence that CPTSD is associated with greater and broader functional impairments than PTSD alone, particularly in relational domains (Hyland et al., 2021; Vallières eta al., 2018). DSM-5, in contrast, conceptualizes PTSD more broadly with four clusters of symptoms, and requires clinically significant impairment in functioning as part of diagnosis (APA, 2013). Including both ICD-11 and DSM-5 functional impairment scales allows cross-framework comparison, while separating PTSD- and DSO-linked impairments within ICD-11 enables a more nuanced understanding of how specific symptom clusters affect functioning. This approach is particularly relevant for refugee children, where trauma exposure may produce both immediate threat-related dysfunctions and enduring relational disturbances due to prolonged displacement and adversity.

While some studies conceptualize functional impairment merely as a consequence of PTSD (Hyland et al., 2021; Vallières eta al., 2018), this study treats PTSD and functional impairment as distinct but related outcomes of trauma exposure. This distinction is supported conceptually, since PTSD reflects clinical symptoms while functional impairment captures disruptions in daily, social, and academic functioning. Empirically, research has shown that trauma exposure can lead to both psychological symptoms (Betancourt et al., 2012), and functional impairments (Mille, & Rasmussen, 2010), and that impairment can occur independently of PTSD. Methodologically, modeling these as separate outcomes allows for a more comprehensive understanding of trauma’s impact on refugee children and highlights cases where children may not meet PTSD criteria yet still experience significant functional difficulties.

Unlike previous studies that examined functional impairment primarily as a consequence of PTSD symptoms in refugee children (Heeke et al., 2020), the present study investigates whether direct trauma exposure—witnessing, physical, or sexual trauma—predicts multiple forms of functional impairment across both ICD-11 and DSM-5 frameworks. This approach recognizes functional impairment as an independent outcome of trauma, particularly relevant in stateless refugee contexts where children may suffer significant disruption in daily life even without meeting diagnostic thresholds for PTSD or CPTSD.

Beyond psychological and social impairments, trauma has been shown to disrupt language development and communication skills. For example, Aviad et al. (2025) demonstrated that children who experienced maltreatment often present with speech and language difficulties, necessitating trauma-informed approaches in speech and language pathology. This suggests that trauma exposure not only affects emotional regulation and functioning but may also impair linguistic development, which is critical for education, identity, and resilience among refugee children. Moreover, recent work has begun to explain the neurobiological mechanisms underlying these difficulties. Hyter (2024) demonstrates how post-traumatic stress alters brain functioning in ways that compromise social pragmatic communication, including perspective-taking, narrative coherence, and adaptive language use. Such disruptions are especially significant in refugee contexts, where children must navigate multiple linguistic environments. Impairments in language functioning, therefore, may contribute to broader psychosocial vulnerabilities, including identity loss, educational deprivation, and difficulties in integration.

### The Mediating Roles of Family and Teacher Support

Children’s exposure to trauma, particularly witnessing violence, as well as physical and sexual trauma do not always translate directly into PTSD, disturbances in self-organization (DSO), or functional impairment. Support systems within the family and school environments often shape how children process traumatic experiences. Family support has been consistently associated with reduced trauma reactivity (Boge et al.,2020), lower stress and anxiety(Ekmeu et al., 2021), and improved overall psychological adjustment. A recent meta-analysis confirmed that higher levels of family cohesion, communication, and emotional support are linked to lower PTSD symptoms in children, while family conflict predicts greater PTSD severity (Ye et al., 2023). Similarly, family support has been shown to buffer against severe mental health outcomes, such as suicidal ideation, by weakening the link between PTSD and depression (Blessing et al., 2023).

However, the protective role of family support may be limited in contexts of severe trauma. Classic studies, such as Kilpatrick and Williams (1997), found that children who directly witnessed domestic violence exhibited PTSD symptoms regardless of maternal well-being or family functioning. Likewise, evidence from stateless Liberian refugees suggests that family support may not always reduce anxiety or related symptoms (Yarseah et al., 2025). These findings indicate that while family support can act as a mediator, its effectiveness may depend on the severity and nature of trauma exposure. This is particularly relevant for refugee children in Nigeria, where prolonged displacement, disrupted family systems, and socio-economic hardship heighten vulnerability to PTSD and CPTSD.

Beyond the family, teachers also play a critical role in mediating the relationship between trauma and child mental health. Refugee children who experience difficulties in emotional regulation and social relationships often look to teachers not only for academic guidance but also for emotional support (Ladhawala, 2021). When teachers are trained in trauma-sensitive approaches, they can serve as protective agents, reducing the severity of PTSD and functional impairment (Bethell et al., 2019). Gamache Martin Cromer,& Freyd (2010) examined teachers’ perceptions of how childhood abuse and neglect affect students’ learning and classroom behavior. Drawing on survey data from preschool through secondary school teachers, the study found that physical and sexual abuse and emotional neglect were associated with significant difficulties in students’ learning, particularly impairments in reasoning and comprehension skills.

Conversely, teachers who lack trauma-informed training may misinterpret trauma-related behaviors, inadvertently exacerbating psychological distress among trauma-exposed students (Alisic et al., 2012). Rather than providing appropriate guidance and emotional support, untrained teachers may respond in ways that unintentionally compound students’ difficulties. In addition, educators working closely with traumatized children often report emotional strain, including intrusive images, feelings of helplessness, fatigue, and symptoms of secondary traumatic stress (Abraham-Cook, 2012). This dynamic is particularly concerning in educational contexts such as Nigeria, where limited access to specialized trauma-informed training may constrain teachers’ capacity to effectively support refugee children with complex trauma histories.

## Materials and Methods

### Ethical Considerations

Ethical clearance for this study was granted by the Ethical Committee for Research, Development, and Innovation at Ekiti State University (Approval No. ORCID/EKSU/EAC/025). All research procedures were conducted in strict accordance with the ethical principles outlined in the Declaration of Helsinki and institutional guidelines for research with human participants. Parental or guardian informed consent was obtained for all child participants, and assent was secured from each child prior to data collection. Participation was entirely voluntary, and participants were explicitly informed of their right to withdraw from the study at any stage without repercussions. Measures were implemented to ensure the confidentiality, anonymity, and psychological safety of all participants throughout the research process.

### Research Design

This study employed a cross-sectional quantitative design to examine the relationship between trauma exposure and PTSD, Complex PTSD (CPTSD), and functional impairment among stateless Liberian and Sierra Leonean refugee children in Oru Refugee Camp, Nigeria. The design was selected to provide a snapshot of participants’ current psychological functioning in relation to prior trauma experiences.

The study tested the associations between types of trauma exposure—physical, emotional, sexual, and witnessed trauma—and psychological outcomes, with gender, age, and language spoken at home (both Nigerian and parental language) included as moderating variables, and family and teacher social support considered as potential mediators.

Data analysis involved descriptive statistics, correlation, and multiple regression analyses. Mediation and moderation effects were examined using SmartPLS, allowing a detailed investigation of the mechanisms and moderators influencing the relationship between trauma exposure and psychological outcomes. This cross-sectional quantitative framework allowed for a systematic assessment of trauma-related psychopathology among refugee children while accounting for key demographic and behavioral factors that may influence these outcomes.

### Age Eligibility and Developmental Rationale

The study focused on refugee children and adolescents aged 6–17 years residing in Oru Refugee Camp. The lower age threshold of six years was selected to include younger children who could provide meaningful self-report data on trauma exposure, psychosocial functioning, and ICD-11–based PTSD and CPTSD symptoms. While Western research suggests that children around age 7 generally demonstrate sufficient self-reflection, temporal understanding, and consistency in reporting internal states (Varni et al., 2007; De Los Reyes & Kazdin, 2005), these benchmarks may not fully generalize to refugee populations experiencing developmental delays from malnutrition, stunting, and prolonged adversity. The study focused on adolescents because this developmental period involves critical maturation of emotion regulation, self-concept, and cognitive systems, which are particularly sensitive to cumulative stressors in displaced populations (Silvers, 2022; Miller & Rasmussen, 2010; Betancourt et al., 2017). This age range was selected to capture the developmental variability in trauma response and psychosocial adaptation across childhood and adolescence.”

#### Sample Size Determination

An a priori power analysis was conducted using G*Power 3.1.9.7 (Heinrich-Heine-Universität Düsseldorf, Düsseldorf, Germany; Faul et al., 2009) to determine the minimum sample required for the most complex structural equation in the model. The analysis was based on the maximum number of predictors (10) pointing to a single endogenous construct, a small-to-moderate effect size (f² = 0.05) as observed in the SmartPLS model (Henseler, Ringle, & Sarstedt, 2015; Sarstedt, Ringle, & Hair, 2017), a significance level of α = 0.05, and statistical power of 0.80. G*Power indicated that a minimum of approximately 150 participants would be required to detect such effects. The final sample of 320 participants exceeded this threshold, ensuring adequate statistical power to estimate structural paths reliably and detect small-to-moderate predictor effects.

#### Sample and Sampling Procedure

A purposive sampling strategy with convenience recruitment was used to select participants from the Liberian and Sierra Leonean refugee population residing in the Oru settlement in southwestern Nigeria. This approach was necessary due to restricted access to the population and logistical constraints associated with conducting research in a densely populated refugee settlement.

Participants were children aged 6–17 years who had resided in the settlement for at least six months and were able to communicate in English, a Nigerian language, or their heritage language. Caregiver consent and child assent were obtained prior to participation. Children were excluded if they were younger than 6 or older than 17 years, exhibited severe psychological distress that could interfere with participation, or were experiencing acute medical crises. A total of 320 participants were included in the final sample. Data were collected using standardized measures and sociodemographic questionnaires, administered individually in a quiet area of the settlement. Research assistants were trained in trauma-informed data collection and read items aloud when necessary to accommodate variations in literacy and language proficiency.

Purposive convenience sampling was justified given the unique characteristics of the Oru refugee settlement, including limited access, high population density, and variability in language and literacy. Random sampling was not feasible under these conditions. This approach ensured that participants who could provide reliable responses and meet inclusion criteria were included, thereby maintaining ethical standards and methodological rigor while capturing meaningful data on trauma exposure, PTSD/CPTSD symptoms, and psychosocial factors among refugee children.

### Instruments and Measurement Model Validation

#### 1. Child and Adolescent Trauma Screen (CATS)

The CATS, developed by Sachser et al. (2017), is a self-report instrument designed to assess trauma exposure, PTSD symptoms, and psychosocial functioning in children and adolescents aged 7–17 years. It comprises 15 items assessing potentially traumatic events, 20 items corresponding to DSM-5 PTSD symptom criteria, and 5 items assessing psychosocial functioning. PTSD items are rated on a four-point Likert scale ranging from 0 (“never”) to 3 (“almost always”), yielding total scores from 0 to 60. A cut-off score of ≥21 is commonly used to indicate clinically significant PTSD symptoms.

Previous validation studies reported excellent internal consistency (α = 0.88–0.94) and strong convergent validity with measures of depression, anxiety, and externalizing behavior (Sachser et al., 2017). In the current sample, the PTSD subscale demonstrated good internal consistency (α = 0.83), strong composite reliability (CR = 0.87), and adequate convergent validity (AVE = 0.56), exceeding commonly recommended thresholds (Hair et al., 2021). These results indicate that the CATS reliably assessed trauma exposure and PTSD symptoms in the refugee children studied.

#### 2. International Trauma Questionnaire – Child/Adolescent Version (ITQ-CA)

The ITQ-CA, developed by Cloitre et al. (2018), is a 22-item self-report instrument assessing ICD-11 PTSD and Complex PTSD (CPTSD) in children aged 7–17 years. It includes six PTSD items covering re-experiencing, avoidance, and sense of current threat, six items measuring disturbances in self-organization (DSO: affective dysregulation, negative self-concept, interpersonal difficulties), and additional items assessing functional impairment across social, academic, and family domains.

Items are rated on a five-point Likert scale from 0 (“never”) to 4 (“almost always”), allowing both categorical and dimensional assessment. The ITQ-CA has been shown to demonstrate excellent reliability and validity for assessing PTSD and Disturbances in Self-Organization (DSO) in children and adolescents, supporting its use as a robust instrument for evaluating ICD-11 PTSD and complex PTSD symptoms (Sigurðardóttir et al., 2025; Haselgruber et al., 2020a). In the current study, the ITQ-CA subscales showed good reliability: PTSD (α = 0.84, CR = 0.87, AVE = 0.45), DSO (α = 0.85, CR = 0.88, AVE = 0.57), and functional impairment (α = 0.83, CR = 0.86, AVE = 0.54). Although the PTSD factor’s AVE was slightly below 0.50, the satisfactory CR and theoretical grounding justified retaining the construct. HTMT ratios were below 0.85, confirming discriminant validity. These findings indicate that the ITQ-CA reliably measured ICD-11 PTSD and CPTSD symptoms in this sample.

#### 3. Child PTSD Symptom Scale – DSM-5 Self-Report (CPSS-SR-5)

The CPSS-SR-5, developed by Foa et al. (2016), is a 20-item self-report measure assessing DSM-5 PTSD symptom severity in children and adolescents. Items are rated from 0 (“not at all”) to 4 (“6 or more times per week / severe”), yielding total scores from 0 to 80, with severity categorized as minimal (0–10), mild (11–20), moderate (21–40), severe (41–60), and very severe (61–80).

Previous studies reported excellent internal consistency (α = 0.92), good test-retest reliability (r = 0.80), and strong convergent and discriminant validity. In the current sample, the CPSS-SR-5 demonstrated acceptable internal consistency (α = 0.77), composite reliability (CR = 0.82), and AVE = 0.45. Despite the AVE being slightly below the 0.50 threshold, the scale’s established validity and satisfactory reliability supported its use for assessing DSM-5 PTSD symptom severity in this population.

#### 4. Social Support Scale for Children (SSSC)

The SSSC, developed by Harter (1985), is a 24-item self-report measure assessing perceived social support from parents, teachers, classmates, and close friends, with six items per subscale. In the present study, only the parent and teacher subscales were administered to align with the social ecological framework.

Items are presented in a structured alternative format, where children select between two statements and indicate whether the chosen statement is “really true for me” or “sort of true for me.” Responses are scored on a four-point scale, with subscale scores ranging from 6–24. Previous studies reported good internal consistency for parent (α ≈ 0.77) and teacher (α ≈ 0.75) subscales (George & Mallery). In the current sample, the parent subscale showed α = 0.78, CR = 0.84, AVE = 0.52, and the teacher subscale showed α = 0.76, CR = 0.85, AVE = 0.54, confirming adequate reliability and convergent validity.

**Table 1:** Analytically Relevant Background Characteristics of Refugee Children (N = 320)

| Analytical Domain | Variable | Category / Statistic | n | % |
| --- | --- | --- | --- | --- |
| Individual Characteristics | Category | n |  | % |
| Gender | Male | 170 |  | 53.1 |
|  | Female | 150 |  | 46.9 |
| Age (years) | Mean (SD) | 12.0 (2.91) |  | — |
| Developmental Stage | 7–10 | 110 |  | 34.4 |
|  | 11–13 | 100 |  | 31.3 |
|  | 14–17 | 110 |  | 34.4 |
| Displacement Background | Liberia | 180 |  | 56.3 |
|  | Sierra Leone | 140 |  | 43.8 |
| Educational Context | Primary | 135 |  | 42.2 |
|  | Secondary | 185 |  | 57.8 |

Demographic variables were organized according to their analytic relevance as individual-, developmental-, and contextual-level controls within the social ecological trauma framework guiding the study. The sample showed balanced representation across gender and developmental stages, supporting the inclusion of these variables as covariates in subsequent analyses.

Table 2 presents the distribution of Nigerian language background across age groups. All children aged 6–8 years (100%) reported speaking a Nigerian language. High proportions were also observed among children aged 11–12 years (87%), 13–14 years (79%), and 15–17 years (74%). The lowest proportion was recorded among children aged 9–10 years, where 71% reported speaking a Nigerian language. Overall, 79% of the sample reported speaking a Nigerian language, indicating that Nigerian languages are widely spoken across all age groups in the study population.

**Table 2:** Nigerian Language Background by Age.

| Age Group (yrs) | Speaks Nigerian Language n (%) | Total (n) |
| --- | --- | --- |
| 6–8 | 28 (100%) | 28 |
| 9–10 | 29 (71%) | 41 |
| 11–12 | 45 (87%) | 52 |
| 13–14 | 60 (79%) | 76 |
| 15–17 | 89 (74%) | 121 |
| Total | 251 (79%) | 318 |

Table 3 presents the distribution of parental home language use among participants. English was the most frequently reported parental home language, reported by 41.0% of households. Indigenous and regional languages were also commonly reported, including Grebo (11.6%), Fula (11.6%), Mandingo (9.4%), and Krun (8.1%). Additionally, 10.6% of parents reported speaking other languages at home. The findings indicate a linguistically diverse home environment, with many children exposed to more than one language.

**Table 3:** Parental Home Language Use (Multiple Responses Allowed)

| Parental Home Language | n (%) |
| --- | --- |
| English | 131 (41.0) |
| Grebo | 37 (11.6) |
| Fula | 37 (11.6) |
| Krun | 26 (8.1) |
| Mandingo | 30 (9.4) |
| Other languages | 34 (10.6) |

**Note.** Percentages are based on the total sample (N = 320). Categories are not mutually exclusive. Fula (Fulfulde) and Mandingo (Mandinka) are transnational languages spoken across several West African countries, including Liberia and Sierra Leone.

### 3.8 Gender Differences in Trauma Exposure

To examine whether there were gender differences in exposure to specific traumatic events, cross-tabulations and chi-square tests were conducted for each trauma item from the Child and Adolescent Trauma Screen (CATS 7–17). Frequencies and percentages for males and females were calculated, and the statistical significance of gender differences was assessed using Pearson’s chi-square test. Effect sizes were reported using Phi (φ) and Cramer’s V coefficients.

**Table 4:** Gender-Specific Trauma Exposure Among Refugee Children (CATS 7–17, N = 320)

| CATS Item | Trauma Type | Trauma Description | Male n (%) | Female n (%) | $\chi^2$ (df=1) | p | $\phi$ |
| --- | --- | --- | --- | --- | --- | --- | --- |
| **CATS1** | Witnessing | Seeing family members slapped, punched, or beaten | 51 (16.1) | 96 (52.5) | 44.60 | <.001 | 0.373 |
| **CATS3** | Witnessing | Seeing community members slapped, punched, or beaten | 134 (42.3) | 123 (67.2) | 19.74 | <.001 | 0.248 |
| **CATS6** | Witnessing | Seeing someone attacked, stabbed, shot at, or hurt badly | 187 (59.1) | 104 (56.8) | 0.17 | .681 | 0.023 |
| **CATS7** | Witnessing | Seeing someone attacked, stabbed, shot at, hurt badly, or killed | 184 (58.4) | 97 (53.0) | 0.92 | .337 | 0.054 |
| **CATS12** | Witnessing | Seeing severe violence in other contexts | 148 (47.4) | 117 (63.9) | 8.69 | .003 | 0.165 |
| **CATS14** | Witnessing | Being around war | 151 (48.2) | 122 (66.7) | 11.05 | .001 | 0.186 |
| **CATS2** | Physical | Serious accident or injury | 94 (68.6) | 87 (47.5) | 14.16 | <.001 | - 0.210 |
| **CATS4** | Physical | Slapped, punched, or beaten in family | 88 (64.2) | 82 (44.8) | 11.87 | .001 | - 0.193 |
| **CATS5** | Physical | Slapped, punched, or beaten by non-family | 86 (62.8) | 85 (46.4) | 8.39 | .004 | - 0.162 |
| **CATS11** | Physical | Attacked, stabbed, shot at, or hurt badly | 87 (63.5) | 79 (43.2) | 12.98 | <.001 | - 0.201 |
| **CATS13** | Physical | Stressful or scary medical procedure | 88 (64.2) | 75 (41.0) | 16.95 | <.001 | - 0.230 |
| **CATS8** | Sexual | Someone older touching private parts | 73 (53.3) | 125 (68.3) | 7.49 | .006 | 0.153 |
| **CATS9** | Sexual | Forced or pressured sexual activity | 70 (51.1) | 132 (72.1) | 14.89 | <.001 | 0.216 |
| **CATS10** | Emotional | Upsetting or scary events | 56 (40.9) | 122 (66.7) | 21.11 | <.001 | 0.257 |
| **CATS15** | Emotional | Other stressful or scary events | 62 (45.3) | 125 (68.3) | 17.14 | <.001 | 0.231 |

**Note:** Percentages are calculated within gender. Negative $\phi$ -values indicate higher prevalence among male children. $\chi^2$ = Pearson chi-square; $\phi$ = Phi effect size for 2×2 tables

Table 5 presents gender-specific patterns of trauma exposure among refugee children. Frequencies and percentages were calculated within gender, and chi-square analyses were conducted for each trauma item, with Phi (φ) coefficients reported as measures of effect size for 2×2 comparisons.

**Table 5.** Trauma Reported as Most Disturbing by Gender (CATS 7–17, N = 320)

| Trauma Category (Most Disturbing) | Male n (%) | Female n (%) | Total n (%) |
| --- | --- | --- | --- |
| **Serious accident or injury** | 26 (19.0) | 42 (23.0) | 68 (21.3) |
| **Slapped / beaten in family** | 24 (17.5) | 18 (9.8) | 42 (13.1) |
| **Attacked / stabbed / hurt badly** | 21 (15.0) | 22 (12.0) | 43 (13.4) |
| **Witnessing family violence** | 14 (10.2) | 27 (14.8) | 41 (12.8) |
| **Witnessing community violence** | 12 (8.8) | 22 (12.0) | 34 (10.6) |
| **Sexual trauma** | 8 (5.8) | 24 (13.1) | 32 (10.0) |
| **Emotional trauma** | 7 (5.1) | 17 (9.3) | 24 (7.5) |
| **War-related events** | 6 (4.4) | 11 (6.0) | 17 (5.3) |
| **Other** | 9 (6.6) | 10 (5.5) | 19 (5.9) |
| **Total** | 137 (100) | 183 (100) | 320 (100) |

**Note:** Percentages are calculated within gender. $\chi^2(14, N = 320) = 30.46, p = .007$ , Cramer's $V = 0.31$ .

Female children reported higher exposure than males across most witnessing, sexual, and emotional trauma items. Statistically significant differences were observed for witnessing family violence (CATS1; 52.5% vs. 16.1%, χ²(1) = 44.60, p < .001, φ = .373), witnessing community violence (CATS3; 67.2% vs. 42.3%, χ²(1) = 19.74, p < .001, φ = .248), severe violence in other contexts (CATS12; 63.9% vs. 47.4%, χ²(1) = 8.69, p = .003, φ = .165), and exposure to war-related events (CATS14; 66.7% vs. 48.2%, χ²(1) = 11.05, p = .001, φ = .186). Female children also reported significantly higher exposure to sexual trauma (CATS8, CATS9; φ = .153–.216) and emotional trauma (CATS10, CATS15; φ = .231–.257).

In contrast, male children reported higher exposure to physical trauma events, including serious accidents or injuries (CATS2; φ = −.210), physical assault within the family (CATS4; φ = −.193), physical assault by non-family members (CATS5; φ = −.162), direct attacks (CATS11; φ = −.201), and stressful medical procedures (CATS13; φ = −.230).

No statistically significant gender differences were observed for certain witnessing items (CATS6, CATS7). Overall, these findings indicate meaningful gender-based differences in trauma exposure patterns, with effect sizes ranging from small to moderate.

A Pearson chi-square test of independence showed a statistically significant association between gender and the type of trauma reported as most disturbing, χ²(14, N = 320) = 30.46, p = .007. The strength of the association was small-to-moderate, as indicated by Cramer’s V = 0.31. Thirteen percent of cells had expected counts less than five, with a minimum expected count of 3.00.

Frequencies and percentages indicated that male children most commonly reported serious accidents or injuries (19.0%), being slapped or beaten in the family (17.5%), and being attacked or hurt badly (15.0%) as the most disturbing trauma. Female children most frequently reported serious accidents or injuries (23.0%), witnessing family or community violence (12–15%), and sexual trauma (13.1%) as the most disturbing events.

Table 6 presents the prevalence of PTSD and CPTSD among the 320 refugee children based on ITQ scoring. PTSD was more prevalent than CPTSD, with half of the participants meeting criteria for PTSD (50.0%; 49.6% of males and 50.3% of females) and 24.1% meeting criteria for CPTSD (23.4% of males and 24.6% of females). Approximately 25.9% of participants did not meet criteria for either diagnosis. These findings highlight that PTSD symptoms were roughly twice as common as CPTSD symptoms in this population, and no meaningful gender differences were observed. Using ITQ makes clear the distinction between PTSD-only and CPTSD, emphasizing that these are separate, clinically relevant diagnostic categories.

**Table 6:** Prevalence of PTSD and CPTSD Diagnoses Among Refugee Children (N = 320) According to the ITQ.

| **Diagnosis** | **Male n (%)** | **Female n (%)** | **Total n (%)** |
| --- | --- | --- | --- |
| **PTSD** | 68 (49.6) | 92 (50.3) | 160 (50.0) |
| **CPTSD** | 32 (23.4) | 45 (24.6) | 77 (24.1) |
| **Subthreshold / No Diagnosis** | 37 (27.0) | 46 (25.1) | 83 (25.9) |
| **Total** | 137 | 183 | 320 |

Note. Percentages for males and females are calculated within each gender; total n (%) reflects the proportion of the full sample. PTSD and CPTSD diagnoses were determined using the International Trauma Questionnaire (ITQ), which distinguishes PTSD-only from CPTSD in accordance with ICD-11 criteria.

As indicated in Table 7, analysis of symptom clusters among the 320 refugee children indicated that trauma-related distress was highly prevalent, though the frequency of specific symptom domains varied. Among PTSD clusters, re-experiencing symptoms were the most common (43.8%), followed closely by avoidance (41.6%) and sense of threat (37.5%). In contrast, disturbances in self-organization (DSO)—the core features of CPTSD—were less frequent, with affective dysregulation reported by 14.1%, negative self-concept by 12.5%, and disturbances in relationships by 10.9% of the sample (Table 12). These findings suggest that while acute posttraumatic symptoms are widespread, fewer children exhibit the enduring emotional and interpersonal difficulties characteristic of CPTSD.

**Table 7:** Prevalence of PTSD and CPTSD Symptom Clusters and Diagnoses by Gender (ITQ Criteria)

| Symptom Cluster / Diagnosis | Male n (%) | Female n (%) | Total n (%) |
| --- | --- | --- | --- |
| **PTSD Clusters** |  |  |  |
| **Re-experiencing** | 60 (43.8) | 80 (43.7) | 140 (43.8) |
| **Avoidance** | 58 (42.3) | 75 (41.0) | 133 (41.6) |
| **Sense of Threat** | 50 (36.5) | 70 (38.3) | 120 (37.5) |
| **PTSD Diagnosis (overall)** | 68 (49.6) | 92 (50.3) | 160 (50.0) |
| **DSO Clusters** |  |  |  |
| **Affect Dysregulation** | 20 (14.6) | 25 (13.7) | 45 (14.1) |
| **Negative Self-Concept** | 18 (13.1) | 22 (12.0) | 40 (12.5) |
| **Disturbances in Relationships** | 15 (10.9) | 20 (10.9) | 35 (10.9) |
| **CPTSD Diagnosis (overall)** | 32 (23.4) | 45 (24.6) | 77 (24.1) |
| **Total Sample** | 137 | 183 | 320 |

**Notes:** Each cluster and diagnosis reflects participants who met both the symptom threshold and functional impairment criteria, in accordance with ITQ scoring standards. Functional impairment is already included in these numbers and should not be listed separately.

Having established the distribution of PTSD and CPTSD symptom clusters, the next step was to examine PTSD Symptom Clusters of the DSM-5 symptom intensity and the degree to which these symptoms interfered with daily life of refugees children in the disbanded Oru refugees camp

### PTSD Symptom Clusters of DSM-5

To further explore symptom severity, mean scores from the *DSM-5 PTSD Symptom Scale* were examined. As shown in Table 8, re-experiencing symptoms had the highest mean (M = 1.75, SD = 1.02), followed by negative alterations in cognition and mood (M = 1.66, SD = 1.08) and arousal/reactivity (M = 1.61, SD = 1.10). Avoidance behaviors were relatively less intense (M = 1.33, SD = 1.14 Functional impairment (M = 0.57, SD = 0.49) indicated that trauma symptoms interfered considerably with daily functioning, including school performance, emotional regulation, and social relationships. Given that trauma exposure and symptom intensity may vary across genders, further analysis examined the distribution of full and partial PTSD diagnoses by gender, as shown in Table 8.

**Table 8:** Descriptive Statistics for PTSD Symptom Clusters (DSM-5 PTSD Scale, N = 320)

| Symptom Cluster | Included Items | M | SD | N |
| --- | --- | --- | --- | --- |
| **Re-experiencing (Cluster B)** | 1–5 | 1.75 | 1.02 | 320 |
| **Avoidance (Cluster C)** | 6–7 | 1.33 | 1.14 | 320 |
| **Negative Alterations in Cognition and Mood (Cluster D)** | 8–14 | 1.66 | 1.08 | 320 |
| **Arousal and Reactivity (Cluster E)** | 15–20 | 1.61 | 1.10 | 320 |
| **Functional Impairment** | B1–B5 | 0.57 | 0.49 | 320 |

When PTSD diagnosis was examined according to DSM-5 criteria, 31.3% of children met the full diagnostic threshold, while 68.7% presented partial PTSD, endorsing one or more clusters without meeting the full criteria (Table 9). Full PTSD was somewhat more common among females (34.4%) than males (27.0%), suggesting that while both genders experienced significant trauma symptoms, girls were slightly more likely to meet the full diagnostic profile.

**Table 9:** Prevalence of Full and Partial DSM-5 PTSD by Gender (N = 320)

| Gender | Partial<br>PTSD n<br>(%) | Full<br>PTSD n<br>(%) | Total<br>(N) |
| --- | --- | --- | --- |
| **Male** | 100 (73.0) | 37 (27.0) | 137 |
| **Female** | 120 (65.6) | 63 (34.4) | 183 |
| **Total** | **220 (68.7)** | **100**<br>**(31.3)** | **320** |

**Note.** Partial PTSD = endorsement of one or more symptom clusters without meeting full DSM-5 criteria. Full PTSD = endorsement of all four clusters and functional impairment.

Overall, PTSD was approximately twice as prevalent as CPTSD in this population of stateless Liberian and Sierra Leonean refugee children. Symptomatically, trauma re-experiencing and avoidance were the most frequent manifestations, while disturbances in self-organization appeared less common. The moderate gender variation—where girls more frequently met full PTSD criteria—likely reflects their greater exposure to witnessing, sexual, and emotional trauma, as noted in the trauma exposure tables.

These findings collectively indicate that while acute posttraumatic symptoms are widespread, the deeper and more enduring disturbances characteristic of CPTSD affect a smaller subset of children, consistent with World Health Organization (ICD-11) distinctions between PTSD and CPTSD and with APA (DSM-5) conceptualizations of trauma-related psychopathology in chronically displaced populations.

### Differentiation between DSM-5 PTSD and Complex PTSD

Findings from the current study indicate that DSM-5 PTSD symptoms were more prevalent than ICD-11 Complex PTSD (CPTSD) symptoms among refugee children in the Oru camp. Specifically, 43.8% met the criterion for re-experiencing, 41.6% for avoidance, and 37.5% for sense of threat, suggesting that trauma-related distress and hyperarousal were widespread. In contrast, fewer children met criteria for the Disturbances in Self-Organization (DSO) clusters, with 14.7% showing affective dysregulation, 12.5% negative self-concept, and 10.9% disturbances in relationships. These results imply that while many children exhibit acute trauma responses consistent with DSM-5 PTSD, only a smaller subset show the enduring self-organization difficulties characteristic of CPTSD. This distinction may reflect differences in trauma type and chronicity, as girls—who experienced more witnessing, sexual, and emotional trauma—may be more vulnerable to developing DSO symptoms, whereas boys’ higher exposure to physical trauma aligns more closely with PTSD symptom expression. To further examine how trauma exposure levels relate to PTSD and CPTSD symptom severity, descriptive analyses were conducted across different trauma types, as presented in Table 9.

Table 10 presents the mean DSM-5 PTSD symptom scores across varying levels of trauma exposure among stateless Liberian and Sierra Leonean refugee children. Across all trauma types, higher exposure levels were associated with greater PTSD severity, demonstrating a consistent dose–response relationship.

**Table 10:** Categorized Trauma Exposure Levels and Mean PTSD (DSM-5) and CPTSD Scores among Refugee Children.

| Trauma Type | Exposure Category | Mean DSM-5 PTSD (SD) | n (%) | Interpretation |
| --- | --- | --- | --- | --- |
| Physical Trauma | Low (0–2) | 2.35 (0.71) | 118 (36.9%) | Lower PTSD severity |
|  | Moderate (3–4) | 3.01 (0.85) | 102 (31.9%) | Moderate PTSD symptoms |
|  | High (5–6) | 3.47 (0.92) | 100 (31.3%) | High trauma–PTSD link |
| Emotional Trauma | Low (0–2) | 2.22 (0.64) | 95 (29.7%) | Lower PTSD severity |
|  | Moderate (3–4) | 2.96 (0.78) | 110 (34.4%) | Moderate PTSD symptoms |
|  | High (5–6) | 3.53 (0.88) | 114<br>(35.9%) | High trauma–PTSD<br>link |
| **Witnessed**<br>**Trauma** | Low (0–2) | 2.40 (0.70) | 148<br>(46.3%) | Lower PTSD severity |
|  | Moderate (3–4) | 3.10 (0.84) | 82<br>(25.6%) | Moderate PTSD<br>symptoms |
|  | High (5–6) | 3.58 (0.95) | 90<br>(28.1%) | High trauma–PTSD<br>link |
| **Sexual Trauma** | None (0) | 2.31 (0.66) | 210<br>(65.6%) | Lower PTSD severity |
| | Present ($\geq 1$) | 3.25 (0.90) | 110<br>(34.4%) | Elevated PTSD<br>symptoms |

**Note:** Exposure categories (Low, Moderate, High) refer to the number of traumatic events experienced by the child. Mean PTSD scores indicate the severity of symptoms associated with each exposure category. Percentages indicate the proportion of the sample within each exposure category, and do not determine the severity label. “High trauma–PTSD link” refers to mean PTSD scores that are elevated relative to other exposure levels.

For physical trauma, children with high exposure (M = 3.47, SD = 0.92) reported markedly higher PTSD symptoms compared to those with low exposure (M = 2.35, SD = 0.71). A similar pattern emerged for emotional trauma, where mean PTSD scores increased from 2.22 (SD = 0.64) in the low group to 3.53 (SD = 0.88) in the high group.

Children who witnessed traumatic events also exhibited a steady rise in DSM-5 PTSD severity, from 2.40 (SD = 0.70) at low exposure to 3.58 (SD = 0.95) at high exposure. Lastly, those with any history of sexual trauma showed substantially higher PTSD means (M = 3.25, SD = 0.90) than those without such experiences (M = 2.31, SD = 0.66).

These results underscore a strong cumulative effect of trauma exposure on PTSD symptom severity, aligning with theoretical expectations that multiple or severe trauma exposures heighten psychological vulnerability among refugee children. To further clarify the relationship between trauma exposure and psychological outcomes, the analysis examined whether different levels of exposure across trauma types were associated with variations in PTSD and CPTSD mean scores.

Table 11 presents mean PTSD (DSM-5) and CPTSD symptom scores across categorized levels of trauma exposure among stateless Liberian and Sierra Leonean refugee children. Across all trauma types—witnessed, physical, emotional, and sexual—there was a clear dose–response pattern, with progressively higher exposure levels associated with increased PTSD and CPTSD symptom burden. For witnessed and physical trauma, increases in mean scores were most pronounced between the moderate and high exposure groups, suggesting that repeated or intense exposure to violence and physical harm substantially exacerbates posttraumatic symptomatology.

Emotional trauma demonstrated a similar graded pattern, with CPTSD scores consistently exceeding PTSD scores across exposure levels, indicating greater sensitivity of disturbances in self-organization (DSO) to emotionally oriented trauma. For example, children with high emotional trauma exposure reported higher CPTSD scores (M = 4.20, SD = 1.15) than those with low exposure (M = 3.22, SD = 0.99). This pattern is consistent with the ICD-11 conceptualization of CPTSD, in which affect dysregulation, negative self-concept, and relational disturbances are particularly responsive to chronic emotional adversity.

In the case of sexual trauma, even low levels of exposure (1–2 events) were associated with notably higher PTSD and CPTSD scores compared to no exposure, underscoring the uniquely severe psychological impact of sexual trauma. Overall, these findings demonstrate a consistent graded association between trauma exposure and posttraumatic symptom burden, supporting a dose–response relationship and highlighting the differentiated sensitivity of CPTSD symptoms to emotional and sexual trauma.

Although mean PTSD and CPTSD scores are reported here, these symptom indices capture clinically relevant disturbances that are closely linked to functional impairment, consistent with the ICD-11 emphasis on impairment in daily functioning as a defining feature of trauma-related disorders.

### Measurement Model Analysis

The measurement model was assessed to establish the reliability and validity of all latent constructs (Table 37). Following the guidelines of Hair et al. (2022) and Henseler, Ringle &, Sarstedt (2015), item reliability, internal consistency, and both convergent and discriminant validity were examined.

### Internal Consistency Reliability

Internal consistency reliability was assessed using composite reliability (CR), which provides a more accurate estimate than Cronbach’s alpha by accounting for variations in indicator loadings and avoiding potential underestimation of reliability (Hair, Ringle &, Sarstedt, 2013; Bagozzi, Yi, 1988; Werts, 1974). Composite reliability values between 0.60 and 0.95 are considered acceptable, with values above 0.70 indicating satisfactory internal consistency (Bagozzi, Yi, 1988; Nunnally JC, Bernstein; 1994). In this study, the CR values for all constructs ranged from 0.86 to 0.90, demonstrating high levels of internal consistency reliability across the measurement model. This indicates that the observed indicators consistently measure their respective latent constructs.

### Convergent Validity

Convergent validity was assessed through the Average Variance Extracted (AVE) for each construct (Fornell & Larcker et al., 1981). An AVE value of 0.50 or higher indicates that a construct explains at least 50% of the variance in its indicators (Bagozzi &, Yi, 1988). As presented in Table 32, all constructs met or exceeded recommended thresholds for convergent validity (Fornell & Larcker, 1981; Hair et al., 2019). Physical Trauma (AVE = 0.690), Witnessing Trauma (AVE = 0.579), and Sexual Trauma (AVE = 0.797) demonstrated strong convergence. Although PTSD/DSM-5 showed a slightly lower AVE (0.449), its composite reliability (CR = 0.866) and Cronbach’s alpha (α = 0.826) exceeded recommended cutoffs (Nunnally & Bernstein, 1994; Hair et al., 2022), indicating adequate convergent validity.

### Discriminant Validity

Discriminant validity was evaluated using the Fornell–Larcker criterion (Fornell & Larcker et al., 1981) and the Heterotrait–Monotrait ratio of correlations (HTMT)(Henseler J, Ringle CM, Sarstedt 2015). Under the Fornell–Larcker criterion, the square root of the average variance extracted (AVE) for each latent construct exceeded its correlations with other constructs, indicating satisfactory discriminant validity.

To corroborate these results, the HTMT ratios were examined (Table 37). All HTMT values were below the conservative threshold of 0.85, and well below the liberal criterion of 0.90, confirming that the constructs are empirically distinct. These results suggest that each latent construct—PTSD/DSM-5, PTSD/WHO, DSO, functional impairment, teacher support, and family support represents a unique conceptual domain with minimal overlap. Collectively, the findings provide strong evidence of discriminant validity and confirm that the measurement model is psychometrically sound and appropriate for subsequent structural analysis.

### Reliability and Validity

Table 12 presents the measurement model results for all constructs included in the study. Internal consistency reliability was assessed using Cronbach’s alpha (α), rho_A (ρA), and composite reliability (CR), with all constructs exceeding the recommended threshold of 0.70, indicating strong internal consistency (Hair et al., 2017). Indicator reliability was evaluated via outer loadings, most of which exceeded the preferred value of 0.70, supporting the adequacy of individual items in representing their respective constructs (Hair, Ringle & Sarstedt, 2013). Convergent validity was assessed using the Average Variance Extracted (AVE), with most constructs demonstrating AVE values above 0.50, indicating that the constructs explain more variance than measurement error (Fornell & Larcker, 1981). The PTSD construct had an AVE of 0.408, slightly below the recommended threshold; however, its high composite reliability (0.872) suggests that the construct remains psychometrically sound for use in structural analyses. Multicollinearity among indicators was examined using the Variance Inflation Factor (VIF), with all values below the conservative threshold of 5, confirming the absence of problematic collinearity (Kock, 2015). Overall, these results demonstrate that the measurement model is both reliable and valid, providing a strong foundation for subsequent structural model analyses.

**Table 12:**
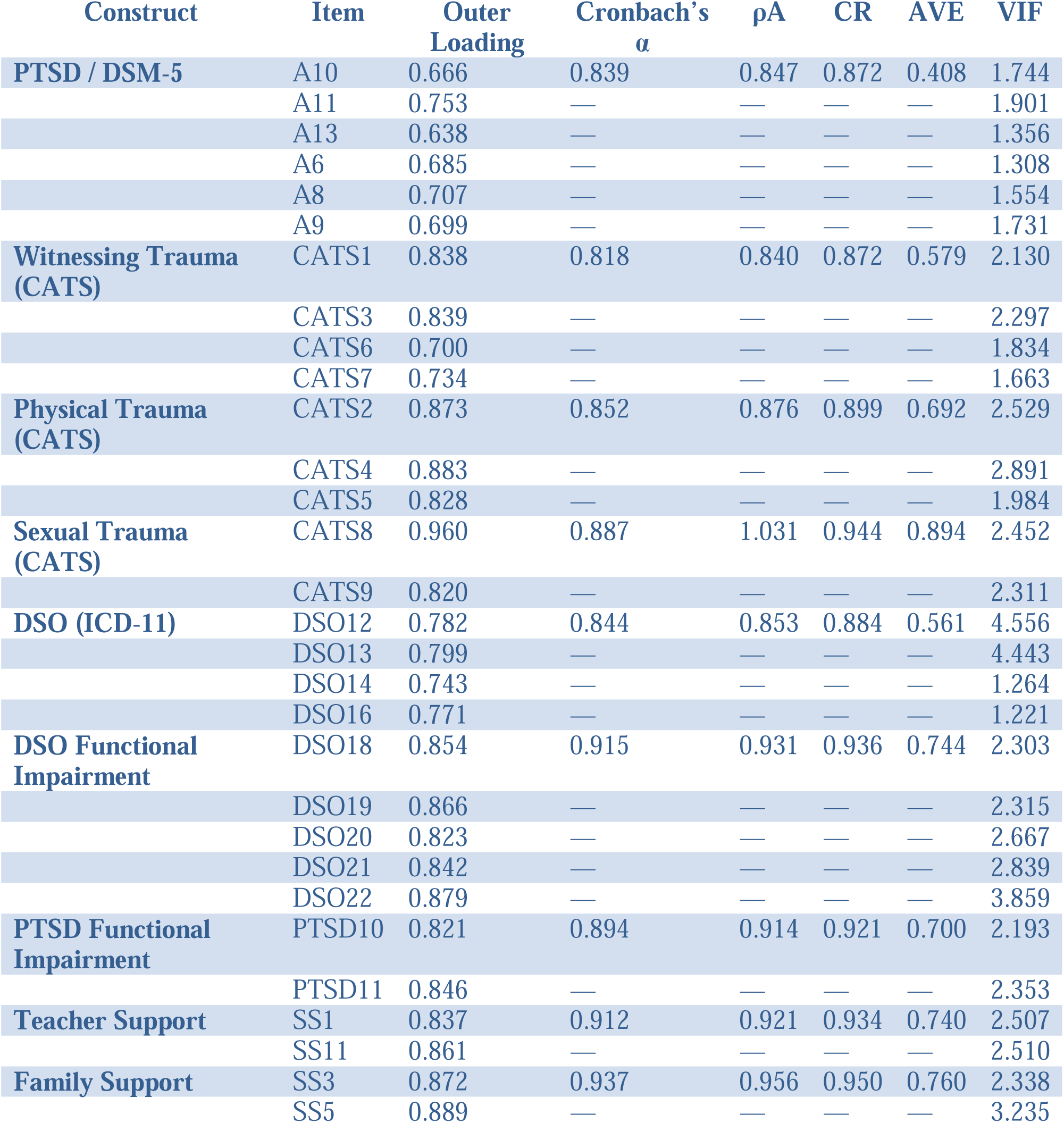
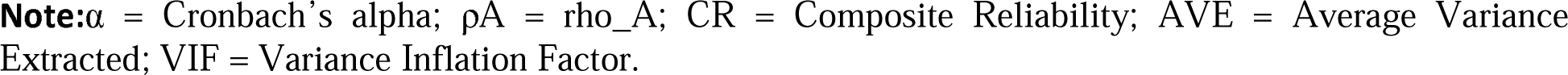
Measurement Model Results (Reliability, Convergent Validity, and Multicollinearity Assessment.

**Table 13:**
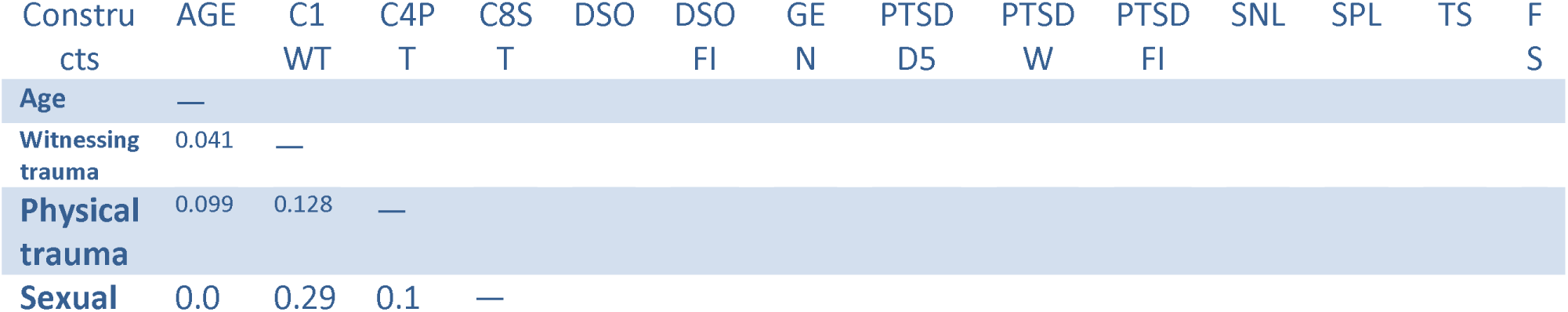
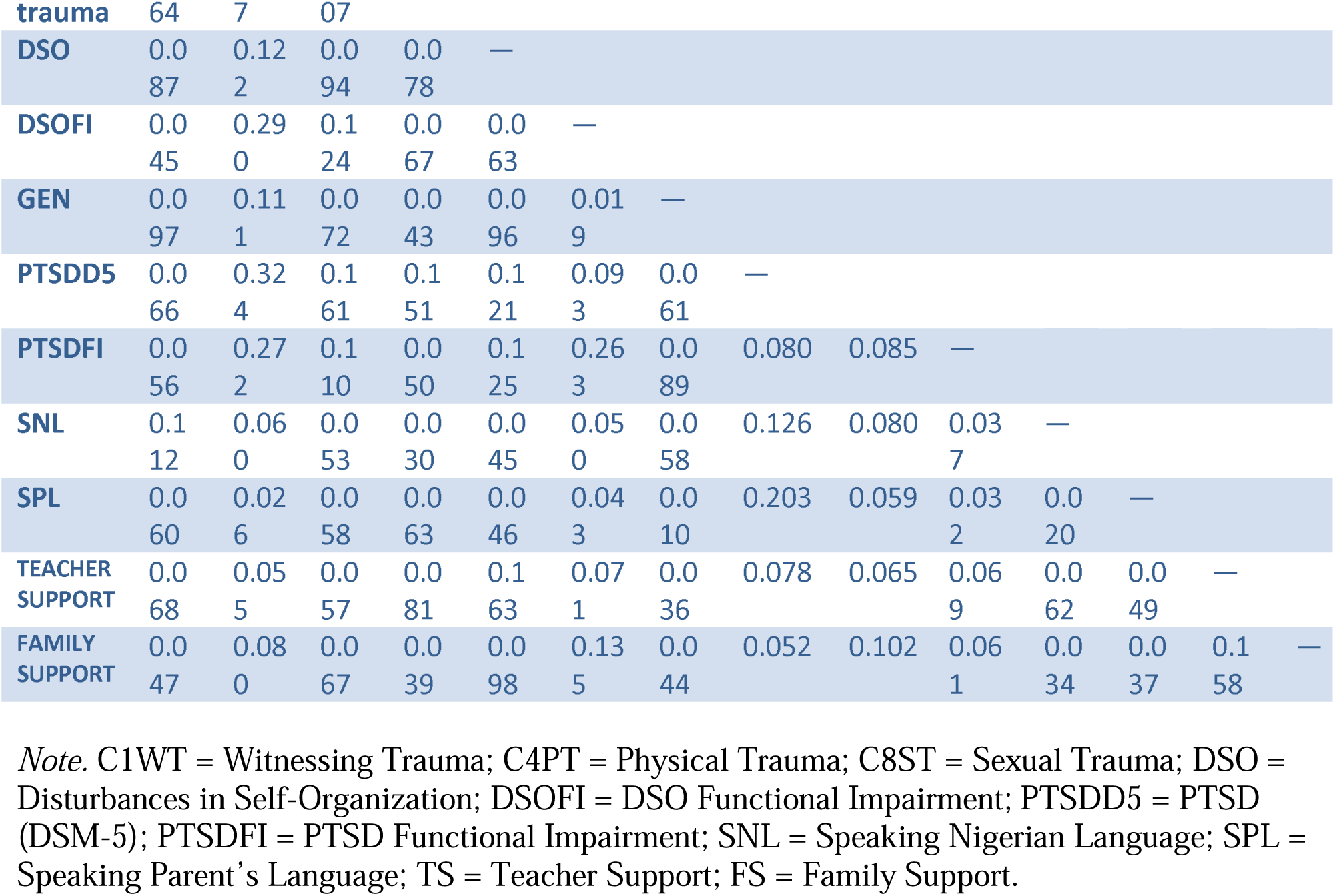
Heterotrait–Monotrait (HTMT) Ratio of Correlations.

Discriminant validity was assessed using the Heterotrait–Monotrait Ratio (HTMT). According to Henseler et al. (2015), HTMT values below 0.85 (strict criterion) or 0.90 (liberal criterion) indicate satisfactory discriminant validity. All values in the present analysis fell below these thresholds, confirming that the constructs are empirically distinct. This demonstrates that each construct—PTSD/DSM-5, DSO, functional impairment, teacher support, and family support—captures a unique conceptual domain with minimal overlap, thereby establishing the discriminant validity of the measurement model.

Table 14 shows the correlation coefficients among study variables ranged from –.15 to .29, indicating low to moderate relationships. The diagonal values (square roots of AVE) were greater than the corresponding off-diagonal correlations, confirming discriminant validity. No correlations exceeded .85, further supporting distinctiveness among constructs (Fornell &, Larcker, 1981).

**Table 14:** Correlation Matrix of Key Study Variables.

| Variable | 1 | 2 | 3 | 4 | 5 | 6 | 7 | 8 | 9 | 10 | 11 | 12 | 13 | 14 |
| --- | --- | --- | --- | --- | --- | --- | --- | --- | --- | --- | --- | --- | --- | --- |
| 1. AGE | **1.00** |  |  |  |  |  |  |  |  |  |  |  |  |  |
| 2. CATS1<br>Witnessing<br>Trauma | .02 | **.76** |  |  |  |  |  |  |  |  |  |  |  |  |
| 3. CATS4 Physical<br>Trauma | -.09 | .06 | **.83** |  |  |  |  |  |  |  |  |  |  |  |
| 4. CATS8 Sexual<br>Trauma | -.07 | .22 | .03 | **.89** |  |  |  |  |  |  |  |  |  |  |
| 5. DSO | .08 | .10 | .07 | .01 | **.78** |  |  |  |  |  |  |  |  |  |
| 6. DSO Functional<br>Impairment | .03 | .26 | .12 | .03 | .04 | **.86** |  |  |  |  |  |  |  |  |
| 7. Gender | .10 | .10 | .07 | .05 | .09 | .02 | **1.00** |  |  |  |  |  |  |  |
| 8. PTSD/DSM-5 | -.04 | .29 | .14 | .13 | .05 | .06 | .03 | **.67** |  |  |  |  |  |  |
| 10. PTSD<br>Functional<br>Impairment | .05 | .25 | .10 | .03 | .11 | .25 | -.09 | .02 | .04 | **.84** |  |  |  |  |
| 11. Speaking<br>Nigerian<br>Language | -.11 | .03 | .05 | .03 | .01 | .05 | -.06 | .12 | .01 | .02 | **1.00** |  |  |  |
| 12. Speaking<br>Parent's<br>Language | .06 | .00 | .06 | .05 | .02 | .04 | .01 | .19 | .01 | .02 | -.02 | **1.00** |  |  |
| 13. Teacher<br>Support | -.07 | .03 | .03 | .06 | .15 | .06 | .01 | .03 | .01 | .06 | .06 | .02 | **.86** |  |
| 14. Family<br>Support | -.05 | .07 | .00 | .01 | .08 | .13 | -.05 | .01 | .11 | .06 | -.02 | -.02 | .15 | **.87** |

**Note.** *N* = [insert sample size]. Values on the diagonal (bold) represent the square roots of the AVE (Fornell–Larcker Criterion). Off-diagonal values represent Pearson correlations between constructs. All correlations are below multicollinearity thresholds ( $r < .85$ ), indicating discriminant validity.

Your structural model examines how different types of trauma—witnessing trauma, physical trauma, and sexual trauma—predict mental-health outcomes among refugee children, specifically PTSD functional impairment, DSO functional impairment, and DSM-5 PTSD symptoms. Family support and teacher support are included as mediating variables to explain *how* trauma exposure translates into these psychological outcomes, reflecting the protective or buffering role of social support systems within the refugee context. In contrast, demographic factors such as age, proficiency in Nigerian languages, and ability to speak the parental/native language function as moderating variables, indicating that the strength of the trauma–outcome relationships may differ depending on children’s developmental stage and linguistic integration.

Gender is included as a control variable to account for baseline differences between boys and girls in both trauma exposure and symptom expression. Overall, the model integrates trauma exposure, social support pathways, demographic moderators, and key psychological outcomes to capture the complex mechanisms influencing PTSD and CPTSD-related impairment among stateless Liberian and Sierra Leonean refugees.

### Structural Model Assessment

Now that we have assessed the measurement model, our next step is to evaluate the structural. The aim was to examine the interrelationships among the key latent constructs such as witnessing trauma, sexual trauma, physical trauma, family and teacher support, functional impairments, DSO and DSM5 as well as other moderating variables.

The model tested both direct effects—such as the effects of witnessing, physical and sexual traumas on functional impairments, DSMD PTSD and DSO. We also tested the indirect effects of family support and teacher support as mediator on the exogenous variables. In addition we addition, we tested sociodemographic variables (speaking Nigerian language and speaking parental language on) as modifiable variables while controlling for age, gender and country of origin Structural relationships were estimated using SmartPLS (version 4.1), a variance-based structural equation modeling technique suitable for complex models and non-normally distributed data (Henseler, Ringle &, Sarstedt, 2015; Sarstedt, Ringle &, Hair, 2017). The model assessment focused on standardized path coefficients (β), statistical significance (p-values), and effect sizes (f²), and explained variance (R² values) for each endogenous variable. This structural evaluation provides insight into the theoretical mechanisms linking experiential avoidance and health-related behaviors with trauma outcomes, while also accounting for the moderating influence of demographic factors. The outer VIF values ranged from 1.31 to 2.89, which are well below the recommended threshold of 5.0 (Hair et al., 2021), indicating that multicollinearity among the indicators was not a concern. Some scholars also propose a more conservative cut-off of 3.3 (Diamantopoulos &, Siguaw, 2006), which was likewise satisfied in this study.

### Model Fit Assessment

Model fit was evaluated using the standardized root mean square residual (SRMR), squared Euclidean distance (d_ULS), geodesic distance (d_G), Chi-square, and the normed fit index (NFI). The SRMR values for the saturated (0.048) and estimated (0.074) models were both below the recommended cut-off of 0.08 (Hu, 1999; Hair et al., 2021), indicating a satisfactory model fit. The NFI values (0.769 and 0.723) were slightly below the ideal threshold of 0.90 but remain acceptable for exploratory PLS-SEM analyses. Overall, these results suggest that the structural model exhibits an adequate level of fit.

As demonstrated in Table 15, global model fit indices indicated that the structural model demonstrated acceptable fit. The standardized root mean square residual (SRMR) value for the estimated model (0.074) was below the recommended threshold of 0.08, indicating a satisfactory approximate model fit in PLS-SEM (Henseler, Ringle &, Sarstedt, 2021; Henseler, Hubona &, Ray, 2015). The discrepancy measures d_ULS (7.466) and d_G (1.337) were close to those obtained for the saturated model, suggesting no substantial model misspecification (Henseler, Hubona &, Ray, 2015).

**Table 15:**
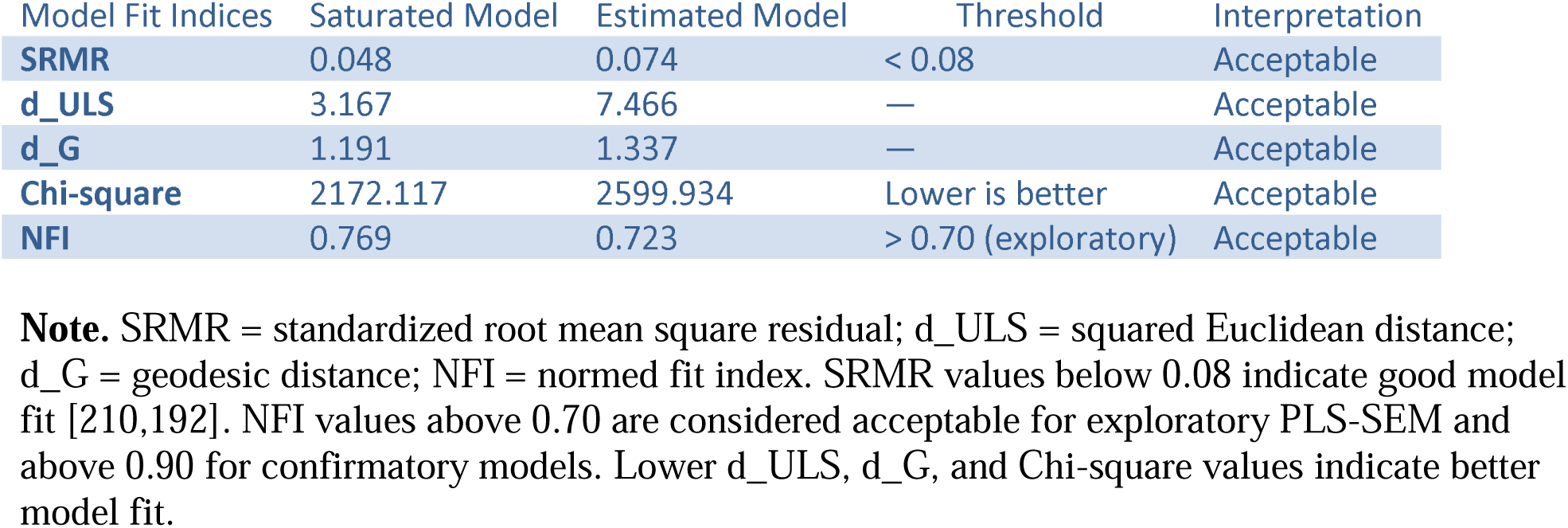
Model Fit.

Although the chi-square value was higher for the estimated model (2599.934) than for the saturated model (2172.117), this pattern is expected in PLS-SEM due to model constraints and sensitivity to sample size, and therefore should not be interpreted as evidence of poor model fit ((Hair, Hult, Ringle &, Sarstedt, 2017; Sarstedt et al., 2016). The normed fit index (NFI) value of 0.723 exceeded the commonly accepted exploratory threshold of 0.70, further supporting acceptable global model fit (Hair, Hult, Ringle &, Sarstedt, 2017). Collectively, these indices indicate that the model exhibits acceptable global fit within the context of PLS-SEM.

**Table 16:** Coefficient of Determination (R²), Adjusted R², and Key Predictors’ Effect Sizes (f²) for Endogenous Constructs.

| Endogenous Construct | $R^2$ | Adjusted $R^2$ | Key Predictors ($f^2$) | Interpretation |
| --- | --- | --- | --- | --- |
| **DSO** | 0.135 | 0.102 | Witnessing Trauma (0.085), Physical Trauma (0.020), Teacher Support (0.028) | Moderate explanatory power; small-to-medium predictor effects |
| **DSO Functional Impairment** | 0.100 | 0.068 | Witnessing Trauma (0.002), Physical Trauma (0.001) | Low explanatory power; trivial effects |
| **PTSD (DSM-5)** | 0.125 | 0.105 | Witnessing Trauma (0.060), Physical Trauma (0.012) | Moderate explanatory power; witnessing trauma strongest predictor |
| **PTSD (WHO)** | 0.018 | 0.002 | Physical Trauma (0.001) | Very low explanatory power |
| **PTSD Functional Impairment** | 0.083 | 0.062 | Witnessing Trauma (0.004) | Weak explanatory power |
| **Teacher Support** | 0.050 | 0.020 | Physical Trauma (0.001), Sexual Trauma (0.001) | Weak explanatory power |
| **Family Support** | 0.013 | -0.003 | Physical Trauma (0.012), Sexual Trauma (0.013) | Weak explanatory power |

**Note.** $R^2$ values represent the proportion of variance explained in each endogenous construct. $f^2$ values of 0.02, 0.15, and 0.35 are typically interpreted as small, medium, and large effects, respectively. Constructs with $R^2$ above 0.10 are considered to demonstrate moderate predictive accuracy in exploratory PLS-SEM models

The coefficient of determination (R²) values indicated that the model explained between 1.8% and 13.5% of the variance across the endogenous constructs. Specifically, DSO (R² = 0.135) and PTSD (DSM-5; R² = 0.125) demonstrated moderate explanatory power, while DSO functional impairment (R² = 0.100), PTSD functional impairment (R² = 0.083), and teacher support (R² = 0.050) showed weak predictive ability.

The effect size (f²) analysis further revealed that witnessing trauma had the most meaningful influence on both DSO (f² = 0.085) and PTSD (DSM-5; f² = 0.060), representing small-to-moderate effects. In contrast, physical and sexual trauma, as well as social support variables, contributed only trivial effects (f² < 0.02).

Collectively, these results suggest that while the model demonstrates statistically valid predictive relationships, the individual predictor effects are modest, highlighting that trauma-related outcomes are influenced by multiple, interacting factors rather than any single dominant predictor.

### Results of Hypothesis Testing

Hypothesis 1a predicted that witnessing trauma would be associated with higher disturbances in self-organization (DSO). This hypothesis was supported, as witnessing trauma significantly predicted higher DSO scores (β = 0.218, 95% CI [0.106, 0.323]). Hypothesis 1b proposed that witnessing trauma would be associated with higher PTSD severity, which was also supported (β = 0.542, 95% CI [0.301, 0.730]). Hypothesis 1c predicted that witnessing trauma would increase PTSD-related functional impairment, and results confirmed this association (β = 0.223, 95% CI [0.129, 0.310]).

Hypothesis 2 stated that physical trauma would predict PTSD severity. This hypothesis was supported, with higher physical trauma associated with higher PTSD symptoms (β = 0.265, 95% CI [0.016, 0.471]). Hypothesis 3 proposed that sexual trauma would predict higher PTSD severity; however, this direct effect was not statistically significant (β = −0.147, 95% CI [−0.375, 0.186]).

#### Summary

Witnessing trauma and physical trauma demonstrated significant direct associations with DSO, PTSD severity, and functional impairment, whereas sexual trauma did not show a significant direct effect on PTSD severity.

### Mediation Analysis

Teacher support exhibited a significant direct negative effect on DSO (β = −0.160, 95% CI [−0.274, −0.033]) and positively predicted family support (β = 0.139, 95% CI [0.021, 0.241]). The indirect effect of teacher support on DSO via family support was statistically significant (β = −0.022, 95% CI [−0.046, −0.006]), indicating partial mediation. The total effect of teacher support on DSO was also significant (β = −0.182, 95% CI [−0.274, −0.033]).

Summary: Teacher support influences children’s self-organization both directly and indirectly through family support, highlighting the protective role of relational contexts in mitigating disturbances in self-organization.

Hypothesis 8 examined whether Nigerian language proficiency moderated the relationship between witnessing trauma and disturbances in self-organization (DSO). The interaction term was significant (β = 0.269, 95% CI [0.067, 0.471]), indicating that the association between witnessing trauma and DSO differed across levels of Nigerian language proficiency. Simple slopes analyses showed that at low levels of proficiency (−1 SD), witnessing trauma was negatively associated with DSO (b ≈ −0.45). At mean proficiency, the association was weaker and close to zero (b ≈ −0.13). At high proficiency (+1 SD), the association became positive (b ≈ +0.12). These results indicate that Nigerian language proficiency alters the magnitude and direction of the trauma–DSO association.

Hypothesis 9 tested whether Nigerian language use moderates the relationship between sexual trauma and disturbances in self-organization (DSO). This hypothesis was supported. The interaction between sexual trauma and Nigerian language use was statistically significant (β = [interaction β], 95% CI [interaction CI]), indicating that the association between sexual trauma and DSO varied across levels of Nigerian language use.

To examine the nature of this interaction, simple slopes analyses were conducted (Figure 3). Among adolescents with low Nigerian language use (−1 SD), sexual trauma was negatively associated with DSO, such that higher levels of sexual trauma corresponded to lower DSO scores (b = [slope_low], 95% CI [CI]). At mean levels of Nigerian language use, the association between sexual trauma and DSO was weak and approximately flat (b = [slope_mean], 95% CI [CI]). In contrast, among adolescents with high Nigerian language use (+1 SD), sexual trauma was positively associated with DSO, with DSO symptoms increasing as sexual trauma exposure increased (b = [slope_high], 95% CI [CI]).

**Figure 2:**
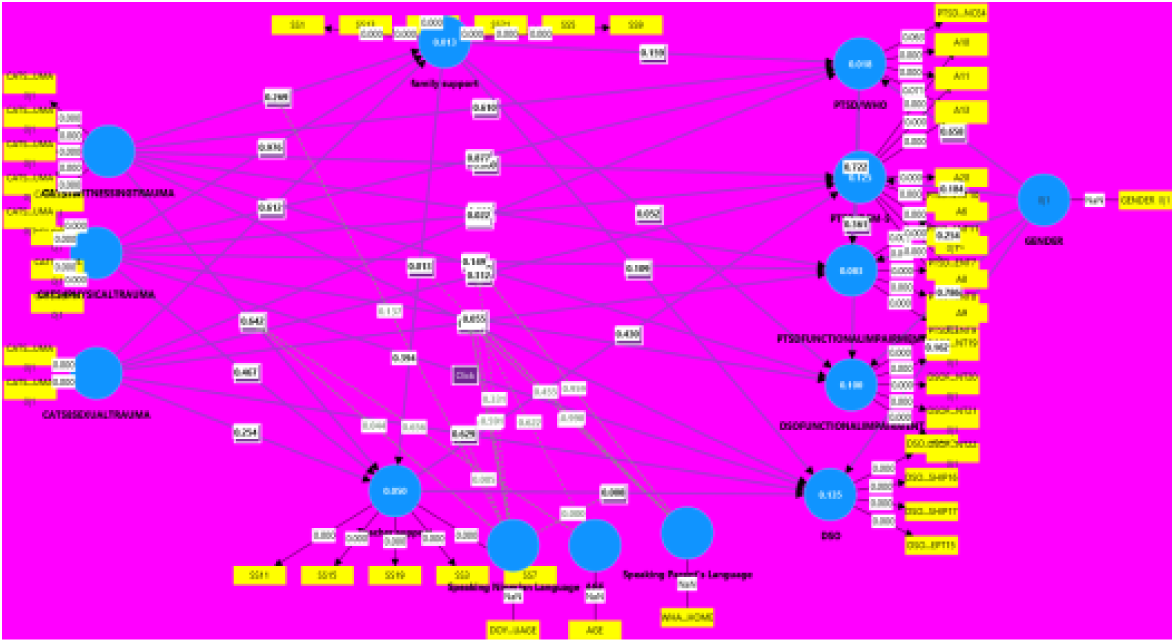
Proposed Structural Model of the Relationships among Study Variables.

**Figure 3.**
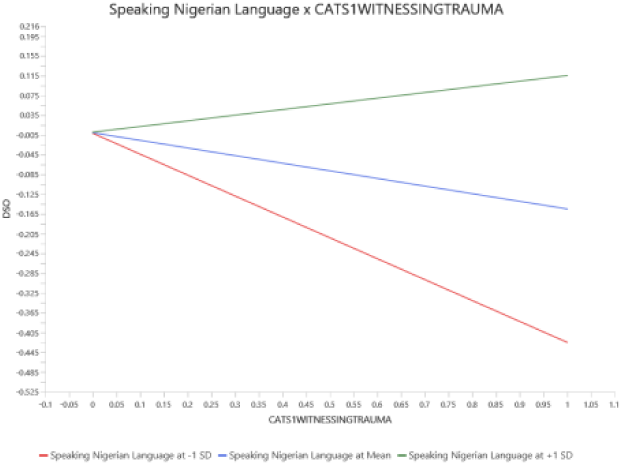
**Note: Figure 3, in**teraction between speaking Nigerian language and witnessing trauma on disturbances in self-organization (DSO). Simple slopes indicate a strong negative association at low language use (–1 SD; b ≈ −0.45), a weaker negative association at mean language use (b ≈ −0.13), and a positive association at high language use (+1 SD; b ≈ +0.12).the **interaction**

Together, these results indicate a crossover interaction, with the direction of the sexual trauma–DSO association reversing across levels of Nigerian language use.

**Figure 4:**
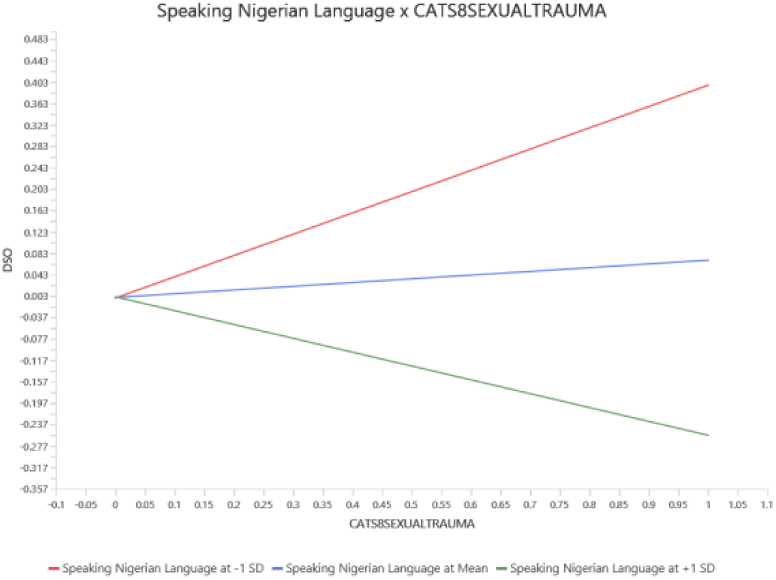
Moderating effect of Nigerian language use on the relationship between sexual trauma and disturbances in self-organization (DSO). Note. Simple slopes are plotted at low (−1 SD), mean, and high (+1 SD) levels of Nigerian language use. Sexual trauma was negatively associated with DSO at low levels of Nigerian language use, showed a weak association at mean levels, and was positively associated with DSO at high levels of Nigerian language use, indicating a crossover interaction.

Hypothesis 10 tested whether Nigerian language proficiency moderated the relationship between teacher support and disturbances in self-organization (DSO). This hypothesis was supported. The interaction between teacher support and Nigerian language proficiency was statistically significant (β = −0.230, 95% CI [−0.338, −0.121]), indicating that the strength of the association between teacher support and DSO varied across levels of language proficiency.

Simple slopes analysis showed that among children with high Nigerian language proficiency (+1 SD), teacher support was strongly and negatively associated with DSO, such that higher levels of support corresponded to substantially lower DSO symptoms. At average proficiency, the negative association was moderate, and among children with low proficiency (−1 SD), the association was weak, suggesting that teacher support was less effective in reducing DSO symptoms at lower levels of language proficiency.

**Figure 5:**
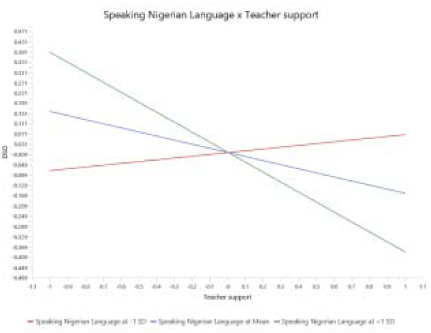
Speaking Nigerian Language × Teacher Support and Disturbances in Self-Organization. **Note.** Simple slopes are plotted at low (−1 SD), mean, and high (+1 SD) levels of Nigerian language proficiency. Teacher support was weakly associated with DSO at low levels of Nigerian language proficiency, moderately associated at mean levels, and strongly negatively associated with DSO at high levels of proficiency, indicating that higher Nigerian languag proficiency amplifies the protective effect of teacher support on DSO symptoms.

Hypothesis 11 tested whether parental language use moderates the relationship between family support and perceived teacher support. This hypothesis was not supported. The interaction term was small and nonsignificant (β = 0.038, 95% CI [–0.060, 0.139]), indicating that the association between family support and perceived teacher support did not differ across levels of parental language use.

In other words, higher or lower parental language engagement did not strengthen or weaken the extent to which family support translated into perceived teacher support. The relationship between family and teacher support appears to be relatively stable, regardless of the language practices within the household.

Hypothesis 12 tested whether parents’ use of their native language moderates the association between witnessing trauma and disturbances in self-organization (DSO). This hypothesis was not supported. The interaction effect was positive but nonsignificant (β = 0.133, 95% CI [–0.084, 0.346]), indicating that parental native-language use did not significantly alter the relationship between witnessing trauma and DSO.

In practical terms, the psychological impact of witnessing trauma on DSO symptoms was comparable across different levels of parental language use. Whether parents frequently used their native language or not, children exposed to witnessing trauma exhibited similar levels of trauma-related disturbances in self-organization.

Hypotheses 13a and 13b tested whether parental language use moderates the relationship between physical trauma and two trauma-related outcomes: PTSD-related functional impairment (H13a) and disturbances in self-organization (DSO) (H13b). Neither hypothesis was supported. The interaction effects were negligible and nonsignificant for both outcomes (H13a: β = −0.002, 95% CI [−0.099, 0.088]; H13b: β = −0.001, 95% CI [−0.095, 0.090]).

These results indicate that the impact of physical trauma on both functional impairment and disturbances in self-organization did not differ across levels of parental language proficiency. In other words, higher parental language use did not attenuate (or exacerbate) trauma-related functional or self-organizational disruptions.

## Discussion

This study examined the impact of different trauma exposures—including witnessing, physical, emotional, and sexual trauma—on PTSD symptoms, disturbances in self-organization (DSO), and functional impairment among Liberian and Sierra Leonean stateless refugee children, while also considering social support and language proficiency. Guided by a social-ecological framework, the analysis explored how individual trauma experiences interact with interpersonal and institutional factors, such as family and teacher support and child Nigerian language use, to shape trauma-related outcomes. Trauma was widespread: males most frequently reported serious accidents (19.0%) and physical abuse (17.5%), whereas females reported serious accidents (23.0%), witnessing family or community violence (12–15%), and sexual trauma (13.1%). Overall, 50.0% of children met ICD-11 PTSD criteria, 24.1% met CPTSD criteria, and 31.3% met full DSM-5 PTSD criteria. Re-experiencing (43.8%) and avoidance (41.6%) were the most common PTSD symptom clusters, while DSO clusters, though less frequent, remained significant (10.9–14.1%). PTSD severity increased with higher exposure to physical, emotional, and witnessed trauma, underscoring the cumulative impact of multiple traumatic experiences on refugee children’s mental health.

H1a stated that witnessing trauma significantly predicted higher DSO functional impairment and it was supported in this current study (β = 0.218, 95% CI [0.106, 0.323]). This suggests that trauma exposure disrupts children’s daily social, emotional, and behavioral functioning—affecting their ability to manage routines, interpersonal relationships, and self-regulation. This aligns with research showing that chronic exposure to violence undermines adaptive functioning in displaced children by overwhelming their coping systems and altering stress-response pathways (Mazrui &, Mazrui, 1998; Singler J. Standards, 2012). Given that DSO is central to CPTSD, these findings indicate that witnessing violence contributes not only to clinical symptomatology but also to broader patterns of psychosocial impairment that shape daily life in the camp.

Also hypothesis 1b stated that witnessing trauma is associated with PTSD/DSM-5 symptoms. The finding of this study demonstrated that **w**itnessing trauma has strongest effect in this hypothesis set, predicting significantly higher PTSD severity (β = 0.542, 95% CI [0.301, 0.730]). This aligns with DSM-5 criteria, which explicitly classify witnessing violence particularly toward caregivers or community members as a qualifying traumatic event capable of producing severe intrusion, avoidance, and arousal symptoms. Developmental trauma literature further shows that young children are especially vulnerable to witnessed trauma, as they lack the cognitive capacity to differentiate danger from safety and rely heavily on caregivers for emotional regulation (APA, 2013). The magnitude of this effect is consistent with studies documenting elevated PTSD symptoms among war-exposed refugee children who witness killings, assaults, or threats within their microsystems (Anderson et al., 2004; Khamis, 2019; Betancourt et al., 2012; Fazel et al., 2012).

Also, Hypothesis 1c stated that witnessing trauma → PTSD functional impairment (children). Witnessing trauma also significantly predicted PTSD-related functional impairment in children (β = 0.223, 95% CI [0.129, 0.310]), reflecting difficulties in school performance, social relationships, emotion regulation, and daily responsibilities. This supports DSM-5 developmental criteria, which require functional impairment as a diagnostic indicator in child PTSD. Literature consistently shows that children exposed to violence exhibit impairments in attention, learning, and emotional stability that interfere with normal developmental trajectories. This aligns with evidence that trauma disrupts neurocognitive and social developmental processes critical for childhood functioning.

Putting hypothesis 1a-c together, the exceptionally strong effect of witnessing trauma observed in this study must be interpreted within the structurally degraded and institutionally abandoned context of the Oru refugee camp. Critical refugee scholarship emphasizes that displacement and statelessness are not merely legal or migratory conditions but are actively produced through state withdrawal, humanitarian disengagement, and institutional neglect, resulting in the erosion of agency, voice, and social recognition over time (Soguk, 1999). Within such contexts, violence becomes normalized and insufficiently regulated, fundamentally reshaping children’s developmental ecologies. Witnessing trauma therefore reflects not only exposure to violent acts but also prolonged immersion in environments characterized by weakened protection systems and fractured adult authority.

Trauma literature demonstrates that witnessing violence—particularly toward caregivers or community members—constitutes a severe form of developmental trauma, capable of producing PTSD symptoms and functional impairment comparable to, and sometimes exceeding, direct victimization (Fazel et al., 2012; Khamis, 2019; Betancourt et al., 2012). In stateless camp settings, the psychological impact of witnessed violence is further amplified by the absence of buffering institutions such as stable schooling, effective child protection services, and consistent humanitarian oversight. This social ecological configuration transforms witnessing trauma into a chronic and cumulative stressor, disrupting emotion regulation, threat appraisal, and meaning-making processes essential for healthy development.

Consistent with social ecological models of trauma exposure, the present findings suggest that witnessing trauma operates as a central mechanism through which structural abandonment and interpersonal violence converge to produce elevated PTSD severity and functional impairment among refugee children. Trauma exposure in this context cannot be understood as discrete events but as embedded within interlocking structural (policy neglect and camp closure), institutional (withdrawal of services), interpersonal (family stress and violence), and intrapersonal (psychological distress) conditions. This explains why witnessing trauma emerged as the strongest predictor across PTSD outcomes in this study, underscoring its role as a core pathway of trauma production in protracted displacement environments.

Hypothesis 2 predicted a direct and significant relationship between physical trauma and PTSD/DSM-5 symptoms among Sierra Leonean and Liberian refugee children in Oru refugee camp. This hypothesis was supported. The results showed that physical trauma had a significant positive effect on PTSD symptoms (β = 0.265, 95% CI [0.016, 0.471]), indicating that children who experienced direct bodily harm reported higher levels of PTSD severity.

This finding is consistent with emerging global evidence on war-affected youth. Research from the ongoing conflict in Ukraine demonstrates that adolescents exposed to combat and displacement exhibit elevated rates of PTSD, anxiety, and depression, with symptom severity highest among those living closest to active conflict zones (Osokina et al., 2023; Badanta et al., 2024). These studies highlight that direct physical harm whether through beatings, assaults, or conflict-related violence is a strong predictor of trauma-related psychopathology, mirroring the pattern observed in the current sample.

Demographic analyses from this study further illuminate the mechanism behind this relationship. Male refugee children reported significantly higher exposure to nearly all indicators of physical trauma. For instance, 64.2% of males (vs. 44.8% of females) reported being slapped, punched, or beaten within the family, χ²(1) = 11.87, p = .001, φ = –.193. Similarly, males were more likely to report physical violence by individuals outside the family (62.8% vs. 46.4%), χ²(1) = 8.39, p = .004, φ = –.162. They also reported higher rates of serious accidents or injuries, being attacked or hurt badly, and undergoing distressing medical procedures. These gendered patterns of exposure suggest that boys in the camp may face unique vulnerabilities to physical victimization, which likely contributes to the observed increase in PTSD symptoms.

Importantly, from a social ecological trauma perspective, these exposures do not occur in isolation but are shaped by the conditions of everyday life in Oru refugee camp. Prolonged displacement, constrained supervision, limited child protection mechanisms, and overcrowded living environments create contexts in which physical violence becomes a recurrent risk embedded within family and community relations. Trauma, therefore, emerges not solely from individual acts of harm but from social and institutional environments that systematically expose children to bodily danger

The broader literature reinforces this interpretation. War-related displacement frequently exposes children to multiple forms of physical danger including assault, exploitation, and chronic community violence all of which are strongly associated with PTSD and impaired emotional regulation (de Alencar Rodrigues et al., 2022). The experiences of Sierra Leonean and Liberian children in Oru Camp therefore align with global patterns documenting the profound psychological toll of physical trauma on conflict-affected youth.

Taken together, these converging lines of evidence underscore that direct bodily harm is a particularly potent predictor of PTSD symptoms in refugee populations. The findings highlight the urgent need for trauma-informed, gender-sensitive interventions that address both the psychological consequences of physical trauma and the structural conditions that place children—especially boys—at heightened risk of harm.

Hypothesis 3 posited that sexual trauma would significantly predict PTSD symptoms among refugee children; however, the effect was non-significant (β = –0.147, 95% CI [–0.375, 0.186]). Demographic analyses revealed that female children in Oru Camp reported significantly higher exposure to sexual trauma, including inappropriate touching (68.3% vs. 53.3% for males, χ²(1) = 7.49, p = .006) and forced sexual activity (72.1% vs. 51.1%, χ²(1) = 14.89, p < .001). Despite this high prevalence, PTSD symptom levels were not directly associated with sexual trauma, suggesting that psychological outcomes are shaped by broader ecological and social factors rather than exposure alone.

Globally, sexual violence against children remains widespread. The 2025 Annual Report on Children and Armed Conflict documented 1,982 verified cases of rape and sexual abuse among children, with the highest numbers reported in Haiti, Nigeria, the Democratic Republic of the Congo, Somalia, and the Central African Republic (Office of the Special Representative of the Secretary-General for Children and Armed Conflict, 2025). This highlights that sexual trauma is both pervasive and contextually embedded in systemic risks associated with conflict, displacement, and weak protective systems.

The observed null association between sexual trauma and PTSD in this study may reflect the dynamic interplay of protective factors and resilience in the children’s immediate ecology. Supportive caregivers, peer relationships, and adaptive coping strategies can buffer the psychological impact of trauma (Rutter, 1987; Garmezy, 1991; Luthar, 2006). Prior research further demonstrates that the psychological consequences of child sexual abuse are strongly contingent on disclosure processes and social reactions. Intrafamilial abuse and disclosure in coercive or unsupportive environments are associated with elevated PTSD risk, whereas delayed or protective disclosure can mitigate symptom severity (Redican et al., 2022; Badanta et al., 2024).

Taken together, these findings underscore the importance of considering sexual trauma within its social and ecological context, including the presence of resilience resources, disclosure dynamics, and the structural vulnerabilities of displaced populations. The results highlight the need for trauma-informed, gender-sensitive interventions that address not only exposure but also the social conditions that shape the psychological impact of sexual violence among refugee children.

Hypothesis four, stated that the impact of teacher support on disturbances in self-organization (DSO) among refugee children occurs indirectly through family support. The results partially supported this hypothesis. Teacher support showed a significant direct negative effect on DSO (β = –0.160, 95% CI [–0.274, –0.033]), indicating that children who received higher levels of teacher support experienced fewer difficulties with emotion regulation, negative self-concept, and interpersonal functioning. However, the indirect pathway through family support was not significant, suggesting that family support did not mediate the relationship between teacher support and DSO.

This pattern is consistent with literature showing that the protective capacity of families can be weakened in contexts of prolonged displacement and high trauma. Classic work by Kilpatrick and Williams (1997) demonstrates that children who witness severe violence may exhibit PTSD symptoms regardless of family functioning. More recent evidence among stateless Liberian refugee families similarly shows that family support does not consistently buffer anxiety or trauma-related symptoms when families themselves are overwhelmed by chronic stressors (Yarseah et al., 2025). These findings suggest that in protracted refugee contexts, family systems may be too strained to operate as strong mediating protective factors.

In contrast, the significant direct effect of teacher support on DSO aligns with research emphasizing the unique role teachers play as stabilizing emotional figures for refugee children. Teachers frequently serve as alternative attachment figures, providing co-regulation, predictability, and emotional validation when family systems are disrupted (Ladhawala, 2021). Trauma-informed teacher practices have been shown to reduce PTSD symptoms and functional impairment (Bethell et al., 2019), supporting the current finding that teacher support independently mitigates DSO symptoms even when family support does not mediate that process.

Taken together, the results suggest that in this population, teachers function as primary protective agents, directly reducing emotional and relational difficulties, while the mediating role of family support is diminished by contextual stressors. This highlights the importance of strengthening school-based psychosocial resources for displaced and stateless children.

Hypothesis 5 proposed that teacher support would be positively associated with family support, and this relationship was supported (β = 0.139, 95% CI [0.021, 0.241]). Refugee children perceiving stronger teacher support also reported greater family support, highlighting the interconnected nature of their support systems.

Within the social ecological framework, teachers occupy the institutional level, directly shaping children’s emotional and relational experiences. Effective teacher support provides co-regulation, predictability, and emotional validation, which buffer the effects of intrapersonal trauma and foster adaptive coping. Yet, teacher capacity is influenced by experience, training, and exposure to trauma-exposed students; some teachers report uncertainty and emotional strain when supporting these children (Alisic et al., 2012; Ladhawala, 2021; Hupe & Stevenson, 2019; Essary et al., 2020).

Despite these challenges, teacher support can enhance family support through coordinated home–school engagement. For example, frequent teacher–family communication has been shown to strengthen parental involvement and student engagement (Kraft & Dougherty, 2013). Trauma-sensitive school models further emphasize that teacher and family engagement operate interdependently to support children’s recovery, creating safe and emotionally supportive learning environments (Cole et al., 2009; Trauma and Learning Policy Initiative, 2017).

From a resilience perspective (Masten, 2001; Garmezy, 1991; Rutter, 2006), teacher and family support function as external protective factors that buffer children from the adverse psychological effects of trauma and displacement. The alignment between teacher and family support reflects a network of protective relationships within the child’s social ecology, enhancing emotional regulation, adaptive functioning, and a sense of security — key markers of resilience. Together, these findings indicate that supportive relationships across institutional and family contexts are mutually reinforcing. Teacher support does not act in isolation but contributes to a broader relational ecology, strengthening family engagement and promoting positive adaptation among refugee children. This underscores the importance of trauma-informed training, adequate institutional resources, and structured home–school communication to optimize protective systems within the social ecology of displaced populations.

Hypothesis 6 proposed that family support would mediate the relationship between teacher support and disturbances in self-organization (DSO), such that higher teacher support would enhance family support, which would subsequently reduce DSO symptoms. The results support this mediated pathway: the indirect effect was statistically significant (β = –0.022, 95% CI [–0.046, –0.006]). Although modest in size, the finding indicates that teacher support contributes to lower DSO symptoms partly by strengthening the family support system.

Importantly, this finding both contrasts with and extends prior research. In well-resourced settings such as the Netherlands, teachers have reported uncertainty and difficulty in supporting traumatized children (Darebo et al., 2024), suggesting limits to teachers’ confidence or training even in stable systems. Yet, in the markedly resource-poor and structurally disrupted context of the former Oru camp, teacher support still emerges as a meaningful protective factor. This contrast highlights the heightened significance of teachers in refugee environments where institutional safety nets have collapsed. At the same time, the current result advances previous work that has largely examined teacher and family support only as direct influences on child mental health (Boge et al.,2020; Ladhawala, 2021), hese findings highlight the critical role of teachers and families as protective agents in the mental health of refugee children. Within the social-ecological framework, supportive actors operate across different levels: teachers provide structure and guidance at the school level, while families provide stability and emotional support at the household level. When teachers are trained in trauma-sensitive approaches, they can reduce the severity of PTSD and functional impairment (Bethell et al., 2019). Conversely, teachers who lack trauma-informed training may misinterpret trauma-related behaviors, inadvertently exacerbating psychological distress (Alisic et al., 2012). Rather than functioning in isolation, teacher and family support interact dynamically, forming interconnected resilience systems that buffer children from cumulative trauma risks. Teacher support strengthens family support, which in turn mitigates DSO symptoms, illustrating how protective influences at different social-ecological levels converge to promote recovery. These results support Hypothesis 6, emphasizing that interventions addressing multiple ecological layers—family, school, and community—are likely to be most effective in improving refugee children’s psychological outcomes.

Hypothesis 7 proposed that teacher support would be negatively associated with disturbances in self-organization (DSO) among refugee children, such that higher teacher support predicts lower levels of emotional dysregulation, negative self-concept, and relational difficulties. The results strongly support this hypothesis. The total effect of teacher support on DSO was significant and negative (β = –0.182, 95% CI [–0.274, –0.033 direct + –0.022 indirect]), indicating that children who perceive stronger teacher support exhibit fewer DSO symptoms. This suggests that teacher support functions as a meaningful protective factor both directly and indirectly through its influence on family support which are supported in the literature

From a social ecological trauma perspective, teachers operate at the institutional (mesosystem) level, providing co-regulation, predictability, structured routines, and emotional validation that directly mitigate intrapersonal trauma. These supports help children develop adaptive coping and relational skills, reducing DSO symptoms even when family microsystems are weakened by displacement, chronic stress, or institutional abandonment. Moreover, the findings suggest a dynamic resilience process: teacher support strengthens family support, which in turn contributes to lower DSO, illustrating how protective mechanisms at different ecological levels interact rather than operate in isolation.

However, working with trauma-exposed children places teachers at risk of secondary traumatic stress (STS) and compassion fatigue (CF), especially in under-resourced educational settings (Hupe & Stevenson, 2019; Essary et al., 2020). Such strain may limit their capacity to provide consistent emotional support, potentially diminishing the protective effect of teacher support on children’s intrapersonal functioning. These dynamics highlight the dual importance of supporting both children and educators through trauma-informed training, supervision, and institutional resources, particularly in refugee education contexts where exposure to cumulative adversity is high.

These findings align with prior research demonstrating that teachers trained in trauma-sensitive approaches can serve as stabilizing agents, reducing PTSD symptoms, functional impairment, and emotional difficulties (Bethell et al., 2019; Ladhawala, 2021). Conversely, untrained teachers may misinterpret trauma-related behaviors and inadvertently exacerbate psychological distress among students (Alisic et al., 2012; Abraham-Cook, 2012). The present study adds value by showing that teacher support does not act solely at the level of individual child-teacher interaction but also **reinforces family systems**, highlighting the interdependent nature of ecological supports in mitigating intrapersonal trauma.

Collectively, these results underscore that in refugee ecologies marked by prolonged displacement and disrupted family systems, teachers play a **critical institutional role**, directly reducing DSO and indirectly supporting resilience through family microsystems. This emphasizes the need for comprehensive strategies that simultaneously enhance teacher capacity and family engagement to promote refugee children’s psychological recovery and adaptive functioning.

The current findings from Hypothesis 8 indicate that Nigerian language proficiency moderates the relationship between witnessing trauma and disturbances in self-organization (DSO), with higher proficiency associated with stronger trauma-related DSO symptoms (β = 0.269, 95% CI [0.067, 0.471]). From a social ecological trauma perspective, this effect reflects the interaction between intrapersonal, relational, and institutional factors in the refugee environment (Miller & Rasmussen, 2010; Panter-Brick et al., 2018). Trauma-exposed children often experience early developmental disruptions, including delays in physical, emotional, and social development (Jaycox et al., 2012), and language acquisition (Hamidli, 2023; Horsman, 2000). Such delays can limit children’s ability to communicate effectively, access support from family members and teachers, and engage with institutional resources.

Proficiency in Nigerian languages may increase children’s engagement with host-community institutions, peers, and teachers, but it also exposes them to additional social stressors, such as discrimination, marginalization, and identity challenges. These chronic stressors can compound intrapersonal trauma, amplifying emotional dysregulation, negative self-concept, and relational difficulties captured in DSO. Consequently, language operates as both a developmental resource and a socio-cultural stressor, shaping how trauma manifests and how children navigate relational and institutional systems. Supporting heritage-language continuity alongside host-language acquisition may therefore help mitigate these vulnerabilities, strengthening resilience across multiple ecological levels (Miller & Rasmussen, 2010; Panter-Brick et al., 2018).

Linguistic hierarchy also plays an important role. In Nigeria, local languages often carry social prestige, while the heritage languages of Liberian and Sierra Leonean refugees are marginalized. Children who shift toward Nigerian languages may experience pressure to abandon their native languages, leading to cultural dislocation and weakened family communication. Loss of linguistic connection to home environments undermines identity stability and disrupts protective family processes, thereby intensifying the emotional and interpersonal consequences of witnessing trauma.

Evidence from European studies further supports this interpretation. Research on children with communication disorders shows robust associations with anxiety, depression, and internalising symptoms (Hahn., Snedeker, & Rabagliati, 2015). Similarly, studies of bilingual and minority-language children demonstrate that low-quality, inconsistent, or disrupted input—either in the heritage language (L1) or in the dominant societal language—predicts poorer mental health outcomes. These patterns mirror the experiences of the refugee children in Oru, many of whom face fragmented exposure to both their home languages and Nigerian languages. Such linguistic instability weakens identity formation, strains family cohesion, and reduces access to culturally meaningful coping strategies. As a result, children lack stable linguistic and social anchors to buffer the psychological effects of trauma.

Taken together, these findings demonstrate that Nigerian language use is not a neutral or universally protective factor. Instead, it interacts with the socio-cultural realities of stateless refugee life—discrimination, out-group identity, linguistic hierarchies, and communication challenges—to magnify the developmental consequences of trauma. This moderation effect underscores the need for psychosocial interventions that support not only host-language acquisition but also preservation of heritage languages, culturally grounded identity development, and trauma-informed engagement within Nigerian community settings.

Hypothesis 9 tested whether Nigerian language use moderates the association between sexual trauma and disturbances in self-organization (DSO). This hypothesis was supported. Sexual trauma was strongly associated with elevated DSO symptoms among adolescents with low Nigerian language use, whereas this association weakened at average levels and attenuated—and potentially reversed—at high levels of language use. The negative simple slope observed at high Nigerian language use (β = –0.328, 95% CI [–0.554, –0.105]) suggests a buffering process whereby frequent engagement in Nigerian languages may reduce the translation of sexual trauma exposure into disruptions in self-concept, affect regulation, and relational functioning.

From a social ecological trauma perspective, this moderation pattern underscores that language functions less as an individual attribute and more as a socially embedded contextual resource. Language use structures access to everyday social participation, communicative legitimacy, and symbolic inclusion within peer and community networks. In refugee camp settings—where formal mental health and psychosocial services are often scarce and structural marginalization is pervasive—such culturally embedded practices may play a particularly salient role in shaping trauma outcomes. By facilitating social interaction and reducing communicative exclusion, language use may alter the relational environment through which sexual trauma is processed, thereby attenuating its association with DSO-related symptoms.

Ethnolinguistic Identity Theory (ELIT) provides further theoretical grounding for this moderation effect by conceptualizing language as a situational identity resource through which individuals negotiate belonging, symbolic status, and identity-related insecurity under conditions of social inequality (Giles & Johnson, 2010; Hahn et al., 2016). Empirical evidence demonstrates that members of structurally disadvantaged groups report higher ingroup pride and collective self-esteem when using socially valued or dominant-group languages, likely because such use reduces perceived social devaluation and status insecurity (Hasan-Aslih et al., 2025). In the present study, frequent Nigerian language use may have similarly mitigated identity-based insecurity and negative self-concept—core components of DSO—thereby weakening the association between sexual trauma and disturbances in self-organization.

Evidence from other ethnolinguistically stratified contexts supports this interpretation. For example, among Palestinian citizens of Israel, disadvantaged group members reported higher ingroup pride and collective self-esteem when using the dominant language (Hebrew) rather than their heritage language (Arabic), highlighting how language use can function as a symbolic and relational resource in unequal social environments (Hasan-Aslih et al., 2025). Although this context differs from refugee displacement, it illustrates a shared psychosocial mechanism whereby access to locally legitimate language practices enhances perceived belonging and social value. In the Oru refugee camp, Nigerian languages similarly carry local communicative authority and social legitimacy; their use may enhance participation, reduce marginalization, and support relational continuity, thereby buffering the impact of sexual trauma on DSO symptoms. Importantly, these findings should not be interpreted as suggesting that heritage language loss is benign or desirable. Rather, they indicate that under conditions of displacement and structural exclusion, engagement with locally legitimate and socially valued languages may offer context-specific psychological protection. Conversely, language attrition, restricted communicative access, or semi-lingualism may undermine identity coherence and social integration, increasing vulnerability to DSO-related outcomes. By conceptualizing language as a moderator—rather than a direct predictor—of the trauma–DSO pathway, this study advances a socially grounded understanding of resilience that emphasizes relational and identity-based resources embedded within the social ecology, rather than individual coping mechanisms alone.

Finally, these findings align closely with ICD-11 conceptualizations of disturbances in self-organization, which emphasize persistent disruptions in self-concept, affect regulation, and relational functioning following chronic interpersonal trauma, including sexual violence (Maercker et al., 2013; Cloitre et al., 2018; Cloitre et al., 2020). Sexual trauma poses a particular risk for DSO because it directly undermines bodily autonomy, identity integrity, and interpersonal trust. The observed moderation suggests that Nigerian language use may support self-organization by stabilizing social identity, enabling emotional expression within shared communicative norms, and sustaining relational engagement within the camp ecology. In this sense, language operates not merely as a cultural marker but as a socially embedded regulatory resource shaping post-trauma adaptation.

To anticipate potential concerns, it is important to note that the observed reversal at high levels of Nigerian language use should not be interpreted as evidence that language exposure negates the psychological impact of sexual trauma. Rather, this pattern likely reflects differential contextual conditions under which trauma is embedded and processed. As this study is cross-sectional, causal inferences cannot be drawn, and it is possible that adolescents with greater social integration—and consequently greater language use—also differ in unmeasured relational or environmental resources. Nevertheless, the moderation effect remains theoretically consistent with social ecological and ethnolinguistic identity frameworks, which emphasize how structurally patterned access to communicative legitimacy and belonging can shape trauma-related outcomes without eliminating trauma exposure itself.

Hypothesis 10 predicted that speaking Nigerian languages would moderate the relationship between teacher support and disturbances in self-organization (DSO), and the results confirmed this: children with high Nigerian-language proficiency experienced substantial reductions in DSO with increasing teacher support, whereas children with low proficiency benefited minimally (β = –0.230, 95% CI [–0.338, –0.121]).

This pattern can be understood in light of the linguistic histories and policies of Liberia and Sierra Leone. Liberia’s monolingual English system and limited institutional support for indigenous languages (Richmond, 1983; Leclerc, 2012) have contributed to heritage-language attrition, leaving many refugee children linguistically underprepared to engage fully with teachers. In Sierra Leone, although multiple local languages are recognized, the absence of a unifying national language (Turay, 2019) fosters linguistic fragmentation, weakening cultural continuity and social belonging.

In Nigeria, linguistic displacement is further intensified, as Yoruba and English dominate schooling and daily life, while heritage languages hold little functional value. Reduced exposure to L1 and sociopsychological pressures, such as displacement and statelessness, accelerate language shift, eroding cultural-linguistic identity (Schmid, 2011; Pavlenko, 2004; Kanu, 2008). From a social ecological trauma perspective, Nigerian-language proficiency shapes children’s ability to benefit from institutional supports. Refugee children fluent in local languages can engage more effectively with teachers, strengthening the protective effect against disturbances in self-organization (DSO). Those with limited proficiency face linguistic and cultural barriers, reducing the buffering impact of teacher support. This demonstrates how interpersonal and institutional supports interact with individual-level capacities to influence trauma outcomes in displaced children.

Hypothesis 11 examined whether parental language use would moderate the relationship between family support and perceived teacher support. The interaction was not significant (β = 0.038, 95% CI [–0.060, 0.139]), indicating that parental language engagement did not influence this interpersonal support pathway. From a social ecological perspective, this finding underscores that protective family-level processes are constrained by broader ecological layers. While family support has been consistently associated with improved psychological adjustment, including lower stress, reduced PTSD symptoms, and stronger emotional regulation (Bøge et al., 2020; Ye et al., 2023; Blessing et al., 2023), these effects primarily operate within the microsystem of the home.

Qualitative research among Somali Bantu and Bhutanese refugees demonstrates that language and financial barriers limit parents’ capacity to assist children academically or engage effectively with educational systems, even when family support is strong (Betancourt et al., 2015). Moreover, chronic displacement and cumulative stress can erode the transferability of protective mechanisms across social domains, weakening mesosystem linkages between family and school (Miller & Rasmussen, 2010; Kira et al., 2012). Thus, the nonsignificant moderation likely reflects a decoupling of family and school systems under conditions of institutional exclusion, where linguistic practices at home do not meaningfully alter children’s perceptions of school-based support.

From a psycholinguistic perspective, nested within this broader ecological context, language attrition and semi-lingualism may further explain these findings. Children exposed predominantly to dominant languages such as English or Yoruba may develop weaker proficiency or emotional attachment to their parents’ heritage languages, reducing the likelihood that parental language use enhances cross-system support processes (Schmid, 2011; Pavlenko, 2004; Montrul, 2016). In this context, even when family support is strong, the language spoken at home does not appear to significantly alter how children perceive or benefit from teacher support.

Together, these results highlight that in contexts of linguistic and institutional displacement, supportive family–school interactions may depend more on relational, emotional, and institutional responsiveness than on heritage language transmission alone. While language remains central to identity and cultural continuity, its role as a moderator of this specific interpersonal support pathway appears limited under conditions of widespread language shift and prolonged displacement.

Hypothesis 12 proposed that parents’ use of their native language would moderate the association between witnessing trauma and disturbances in self-organization (DSO). However, the interaction effect was not significant (β = 0.133, 95% CI [–0.084, 0.346]), indicating that parental language use did not alter the impact of witnessing trauma on emotional dysregulation, negative self-concept, or relational difficulties. Rather than undermining the relevance of language, this finding highlights important limits to its protective role under conditions of severe and cumulative trauma.

From a social ecological trauma perspective, this null moderation effect suggests that language cannot function independently of the ecological and experiential conditions in which it is embedded. At the intrapersonal level, DSO reflects profound disruptions in meaning-making, emotional regulation, and relational trust (Chang et al., 2021). When children are repeatedly exposed to traumatic events, particularly in settings marked by insecurity and institutional abandonment, these internal systems may be destabilized in ways that exceed the regulatory capacity of parental language use alone.

This interpretation is strongly supported by phenomenological accounts of trauma, which demonstrate that extreme or prolonged trauma can disrupt not only psychological functioning but also the lived experience of language itself. Ratcliffe’s work(ND) shows that in conditions of severe trauma, language may be experienced as hollow, estranged, or inadequate for articulating internal states, particularly when trust in others and in the social world has been eroded. Such “non-localized breakdowns of trust” fundamentally alter how communication is experienced, limiting language’s capacity to support emotional containment, self-coherence, or relational repair. Under these conditions, parental language—while culturally and emotionally meaningful—may no longer serve as an effective medium for restoring intrapersonal organization.

Empirical developmental research further supports this explanation. Traumatic exposure in early childhood, including witnessing violence, has been consistently associated with broad disruptions across emotional, social, and communicative domains (Jaycox et al., 2012). Accumulating evidence indicates that trauma-related stress can impair language acquisition and expressive competence, resulting in delayed development, reduced verbal fluency, and diminished communicative effectiveness (Horsman, 2000; Hamidli, 2023). These trauma-related impairments may limit children’s ability to benefit from linguistic input, even when parents continue to use their native language consistently, thereby constraining its potential buffering role.

Importantly, this finding does not contradict research highlighting the protective value of language use in other contexts. For example, Whalen et al. (2022) demonstrate that Indigenous language use and revitalization are associated with positive mental and physical health outcomes when embedded within functioning cultural, institutional, and communal ecologies. The present findings instead illustrate the converse condition especially in settings of statelessness, marginalization, and cumulative adversity, where trust, safety, and institutional support are systematically weakened, language alone may lose its symbolic and regulatory power.

Taken together, the nonsignificant moderation effect observed in Hypothesis 12 underscores a key principle of the social ecological trauma framework: protective processes are contingent, relational, and ecologically scaffolded. Parental language use does not operate as a standalone moderator of trauma-related intrapersonal outcomes when broader ecological systems are compromised. Instead, effective protection against disturbances in self-organization likely requires multi-level supports—including stable caregiving relationships, trauma-informed educational environments, and institutional structures that restore safety, trust, and meaning—within which language can regain its protective function.

This interpretation is strongly supported by language-attrition literature. Schmid (2011) argues that when a heritage language loses functional value and institutional support, “linguistic traffic” from L1 diminishes, leading to both active and passive attrition. This scenario is characteristic of displaced and marginalized groups. Furthermore, research on refugees demonstrates that forced migration, statelessness, and the pressure to assimilate into host societies accelerate linguistic shift (Pavlenko, 2004; Montrul, 2016). In such conditions, families often reduce use of their native language to avoid stigma, marginality, or perceived barriers to integration. Over time, the heritage language loses its emotional currency and becomes less effective as a source of identity, belonging, or resilience.

Hypotheses 13a and 13b examined whether parental language use moderates the association between physical trauma and PTSD-related functional impairment (H13a) and disturbances in self-organization (DSO; H13b). Neither moderation effect was statistically significant (H13a: β = −0.002, 95% CI [−0.099, 0.088]; H13b: β = −0.001, 95% CI [−0.095, 0.090]), indicating that the psychological impact of physical trauma did not differ across levels of parental language proficiency.

From a social ecological trauma perspective, these findings suggest that parental language use does not function as a sufficiently proximal protective mechanism to buffer the effects of physical trauma under conditions of prolonged displacement. Physical trauma involves direct exposure to bodily harm and threat to life, which is known to produce highly salient and enduring neurobiological and psychological effects that may be less amenable to contextual or relational buffering processes. PTSD-related functional impairment reflects disruptions in daily functioning, while disturbances in self-organization (DSO) capture enduring patterns of affective dysregulation, negative self-concept, and relational impairment central to ICD-11 conceptualizations of complex trauma responses (Cloitre et al., 2013, 2018).

The absence of moderation is consistent with models emphasizing the role of chronic daily stressors in shaping mental health outcomes among forcibly displaced populations (Miller & Rasmussen, 2010). In refugee ecologies characterized by persistent material deprivation, insecurity, and institutional withdrawal, such stressors may exert more immediate and sustained influence on psychological functioning than family-level protective practices. Under these conditions, caregiving processes—including parental language use—may retain cultural and relational significance but remain insufficient to attenuate trauma-related impairment when broader structural stressors remain unmitigated (Tol et al., 2011; Siriwardhana et al., 2013; Sim et al., 2018).

Importantly, the lack of a moderating effect does not negate the relevance of parental language use for children’s broader developmental or psychosocial outcomes. Rather, it indicates that mitigating the psychological consequences of physical trauma may require interventions operating at institutional, community, and policy levels, including access to trauma-focused mental health services, educational stability, and material security. Consistent with ecological models of resilience, adaptive outcomes are most likely to emerge when protective processes operate across interconnected systems rather than relying on singular familial or cultural resources (Rutter, 1987; Masten, 2014).

### Implications of the study

The findings of this study have several important implications for theory, policy, and practice concerning trauma among stateless refugee children. The high prevalence of DSM-5 PTSD (50.0%) and ICD-11 CPTSD (24.1%), alongside substantial functional impairment, underscores that trauma in this population is not solely an individual clinical issue but a systemic problem shaped by ecological and institutional conditions. Differential associations between trauma types and symptom profiles highlight the conceptual distinction between PTSD and CPTSD, with physical and witnessed trauma more strongly linked to fear-based PTSD symptoms, and emotional and sexual trauma associated with disturbances in self-organization (DSO). *Theoretically, these findings support a differentiated trauma framework in which symptom expression is shaped by both trauma exposure and access to ecological resources*.

Furthermore, the moderating effect of language proficiency on the relationship between teacher support and DSO indicates that institutional protective factors are contingent upon children’s ability to access them linguistically. *This suggests that language functions not only as a communicative resource but as a structural gateway to psychosocial protection*, emphasizing the centrality of language for equitable educational and mental health outcomes in displacement contexts.

### Recommendations

Based on the present findings, several evidence-informed recommendations emerge. First, trauma assessment in refugee and displacement contexts should integrate both DSM-5 PTSD and ICD-11 CPTSD frameworks. The observed associations between emotional and sexual trauma and disturbances in self-organization (DSO) suggest that reliance on fear-based PTSD models alone may lead to under-identification of complex trauma–related impairment, particularly in chronically displaced populations.

Second, schools serving refugee children should be conceptualized as frontline mental health environments. Teacher training programs should incorporate trauma-informed and developmentally sensitive approaches that address CPTSD-related behaviors such as affect dysregulation, relational withdrawal, and negative self-concept, reducing the risk that trauma responses are misinterpreted as disciplinary or motivational problems.

Third, language support should be prioritized not only as an educational resource but as a psychosocial mechanism of protection. Language-bridging programs, bilingual instruction, and linguistically inclusive pedagogical practices may enhance children’s access to supportive teacher relationships, thereby strengthening school-based protective pathways within a social ecological framework.

Finally, intervention models for refugee children exposed to emotional and sexual trauma should extend beyond fear-reduction paradigms to include components targeting relational safety, emotional regulation, and self-concept repair. Approaches grounded in developmental and complex trauma theory may be better suited to addressing the functional impairments associated with prolonged interpersonal trauma.

### Strength and Limitations of the study

A key strength of the present study lies in its integrative contribution to the literature on language and trauma-related mental health outcomes among stateless refugee children. By examining language-related resources alongside disturbances in self-organization (DSO), PTSD symptoms, and functional impairment, this study extends existing trauma research beyond symptom-focused models and highlights the role of linguistic and institutional pathways in shaping psychological adjustment under conditions of prolonged displacement. The application of a social ecological framework further strengthens the study by situating individual trauma responses within broader interpersonal and institutional contexts, an approach that remains underrepresented in refugee mental health research, particularly in low-resource settings.

Despite these strengths, several limitations warrant consideration when interpreting the findings and also point to important directions for future research. First, the cross-sectional design precludes causal inference regarding the relationships between language proficiency, teacher support, and trauma-related outcomes. Future longitudinal studies are needed to examine developmental trajectories of PTSD and CPTSD and to assess the sustained influence of institutional and linguistic access on DSO symptoms over time. Second, reliance on self-report measures may have led to underreporting of trauma exposure, particularly for stigmatized experiences such as sexual violence. Subsequent research should incorporate clinical interviews, observational methods, or multi-informant assessments to enhance measurement validity and improve trauma disclosure.

Although the study adopts a social ecological perspective, structural-level factors—including legal status, duration of displacement, and access to humanitarian or educational services—were not directly assessed, limiting the ability to fully explain macro-level influences on trauma-related distress. Future studies would benefit from explicitly integrating these structural variables to strengthen multi-level models of refugee mental health. Finally, the findings are context-specific to stateless refugee children residing in Nigeria. Replication across diverse refugee contexts and institutional settings is therefore necessary to determine the generalizability of these findings and to assess whether similar language–trauma dynamics operate in other sociopolitical and cultural environments.

## Conclusion

Trauma-related psychopathology among stateless refugee children is both highly prevalent and ecologically patterned. Different trauma types contribute to distinct outcomes: physical and witnessed trauma were strongly associated with PTSD severity and functional impairment, while emotional and sexual trauma conferred heightened risk for disturbances in self-organization (DSO). Teacher support served as a protective factor only when children possessed sufficient language proficiency, highlighting how institutional resources can be unevenly accessible and how language functions as a key determinant of equitable psychosocial outcomes. Integrating trauma exposure, DSM-5 PTSD, ICD-11 CPTSD, functional impairment, and school-based protective factors within a social ecological framework underscores the need for linguistically inclusive, trauma-informed educational and mental health policies. Addressing these multi-level dynamics is critical for promoting recovery, resilience, and long-term psychosocial well-being among stateless refugee children living in contexts of chronic adversity

## Data Availability

The data for this study is deposited at: 10.5281/zenodo.18885950

https://doi.org/10.5281/zenodo.18885950

## Notes

### Competing Interest Statement

The authors have declared no competing interest.

### Funding Statement

This study did not receive any funding

### Author Declarations

Ethical clearance for this study was granted by the Ethical Committee for Research, Development, and Innovation at Ekiti State University (Approval No. ORCID/EKSU/EAC/025).

